# Learned ultrasound segmentation and deformable CT fusion for augmented reality endovascular surgery

**DOI:** 10.64898/2026.07.15.26358084

**Authors:** Tom Dillon, Diego Quevedo Moreno, Brian Ayers, Emma K. Rutherford, Brett Salomon, Boateng Kubi, Jonah Thomas, Ellen T. Roche

## Abstract

Minimally invasive endovascular procedures offer reduced surgical trauma, shorter recovery times, and improved outcomes, but rely on 2D fluoroscopic X-ray imaging, which provides limited depth perception and exposes patients and clinicians to ionizing radiation. Here we present an augmented reality (AR) system that fuses intravascular ultrasound (IVUS) and electromagnetic (EM) position tracking with preoperative computed tomography (CT) to produce an anatomically accurate, deformation-corrected navigational reference. A robotic device performs ECG-gated pullback of the IVUS probe, capturing 4D aortic motion across the cardiac cycle. We introduce a deep learning architecture for extracting vascular lumen boundaries and side-branch orifices from artifact-prone IVUS streams, and a semantically driven non-rigid CT-IVUS fusion pipeline robust to false positive landmarks. We evaluate the platform with trained surgeons in benchtop phantom studies and in vivo ovine models, and demonstrate its application to fenestrated endovascular aneurysm repair (FEVAR). Compared to fluoroscopy alone, AR guidance significantly reduces cannulation time, radiation exposure, and cognitive workload, while improving procedural efficiency and safety.

---

Compared to open surgery, which requires large incisions for direct organ access, endovascular procedures offer shorter recovery times, reduced perioperative morbidity, lower infection risk, and improved patient outcomes^1^. Common endovascular procedures include renal artery stenting to treat atherosclerotic renal artery stenosis, endovascular aneurysm repair (EVAR) for treatment of aortic aneurysms, and renal denervation for management of refractory hypertension^2–4^. A critical step in many aortic procedures is target vessel cannulation (TVC), the selective navigation of a guidewire or catheter into a specific branch artery from within the aortic lumen to deliver a therapeutic device. TVC difficulty is compounded by the inherent limitations of conventional X-ray fluoroscopic imaging, which provides only 2D representations of 3D vascular anatomy, making it challenging to judge the orientation and angulation of branches required for successful cannulation^5^. Studies have shown that TVC accounts for a substantial portion of total procedure time and cumulative radiation dose in EVAR^5,6^, with cannulation times ranging from 5 to 30 minutes per vessel depending on anatomical complexity, vessel tortuosity, and operator experience. Despite the use of lead aprons, surgeons face occupational radiation exposure associated with an elevated incidence of brain tumors and other malignancies^7,8^. Moreover, each cannulation attempt carries risk of unintentional vascular injury, since guidewire manipulation without precise 3D feedback can result in vessel dissection or perforation^9^.

To address the limitations of 2D fluoroscopic imaging, contemporary interventional practice has increasingly employed *image registration* that integrates high-resolution 3D preoperative imaging with intraoperative visualization^10–13^. The most common form of multimodal image registration in endovascular surgery is CTA-to-X-ray registration^14,15^ which fuses preoperative 3D imaging with intraoperative 2D fluoroscopy based on easily identifiable anatomical landmarks such as vertebral bodies or pelvic bone contours. Clinical guidelines suggest that spatial deviation between the registered CT location and the true location of aortic branch orifices should not exceed 3 mm (approximately half the diameter of a typical renal artery) to facilitate reliable TVC^16,17^. However, studies report image registration errors of 4– 10 mm can occur in up to 40% of cases^17,18^. This discrepancy arises from limited depth information from fluoroscopic X-ray and the assumption of rigid body registration, which does not account for potential non-rigid deformation factors of the aorta such as variations in patient posture or disease progression^19^.

These limitations motivate intravascular sensing approaches that localize anatomy directly, rather than inferring it from external imaging. Intravascular ultrasound (IVUS) is an interventional imaging modality that uses a miniature ultrasound transducer at the catheter tip to generate real-time, high-resolution 2D cross-sectional images of blood vessel anatomy, while electromagnetic (EM) tracking and fiber optic shape sensing (FOSS) provide real-time 3D instrument localization without line-of-sight constraints. IVUS coupled with position sensing enables direct visualization of branch orifices from within the vessel lumen, without relying on external landmark assumptions inherent to fluoroscopic registration. Prior work has explored IVUS-EM fusion for 3D aortic reconstruction^20–22^, progressively incorporating CT-driven pose optimization to exploit preoperative priors and denoise EM measurements, which are susceptible to distortion from metallic operating room equipment^23–25^. However, these methods either require fiducial markers or manual initialization, or neglect non-rigid deformation of the aorta, including that from cardiac-driven motion. In subsequent work by Zhao et al^26^, the initialization problem was addressed by jointly optimizing both the global rigid CT-IVUS registration and individual frame poses. This approach still fundamentally assumes that no non-rigid deformation has occurred since the preoperative CT scan.

To build upon existing rigid endovascular registration systems and address the key clinical challenges associated with TVC, we propose an AI-driven augmented reality (AR) platform for navigational guidance in the aorta driven by semantic non-rigid registration of IVUS and CT imaging. The platform integrates several complementary sensing modalities to enable dynamic 4D reconstruction of aortic anatomy as shown in Fig. 1. An IVUS catheter is coupled to an EM position sensor, providing simultaneous imaging of vessel cross-sections and 6 degree-of-freedom pose data of the catheter tip. The hybrid catheter motion is controlled by a robotic pullback device synchronized with the patient’s cardiac cycle via electrocardiogram (ECG) gating, enabling the system to capture and account for cyclic aortic deformation induced by cardiac motion. All intraoperative data (IVUS, EM, and ECG) is fused with preoperative CT imaging on a nearby workstation computer and the resulting dynamic 4D reconstruction is rendered in real time, providing an augmented reality representation of both the anatomy and the position of surgical instruments within it for guiding TVC.

**Fig. 1.**
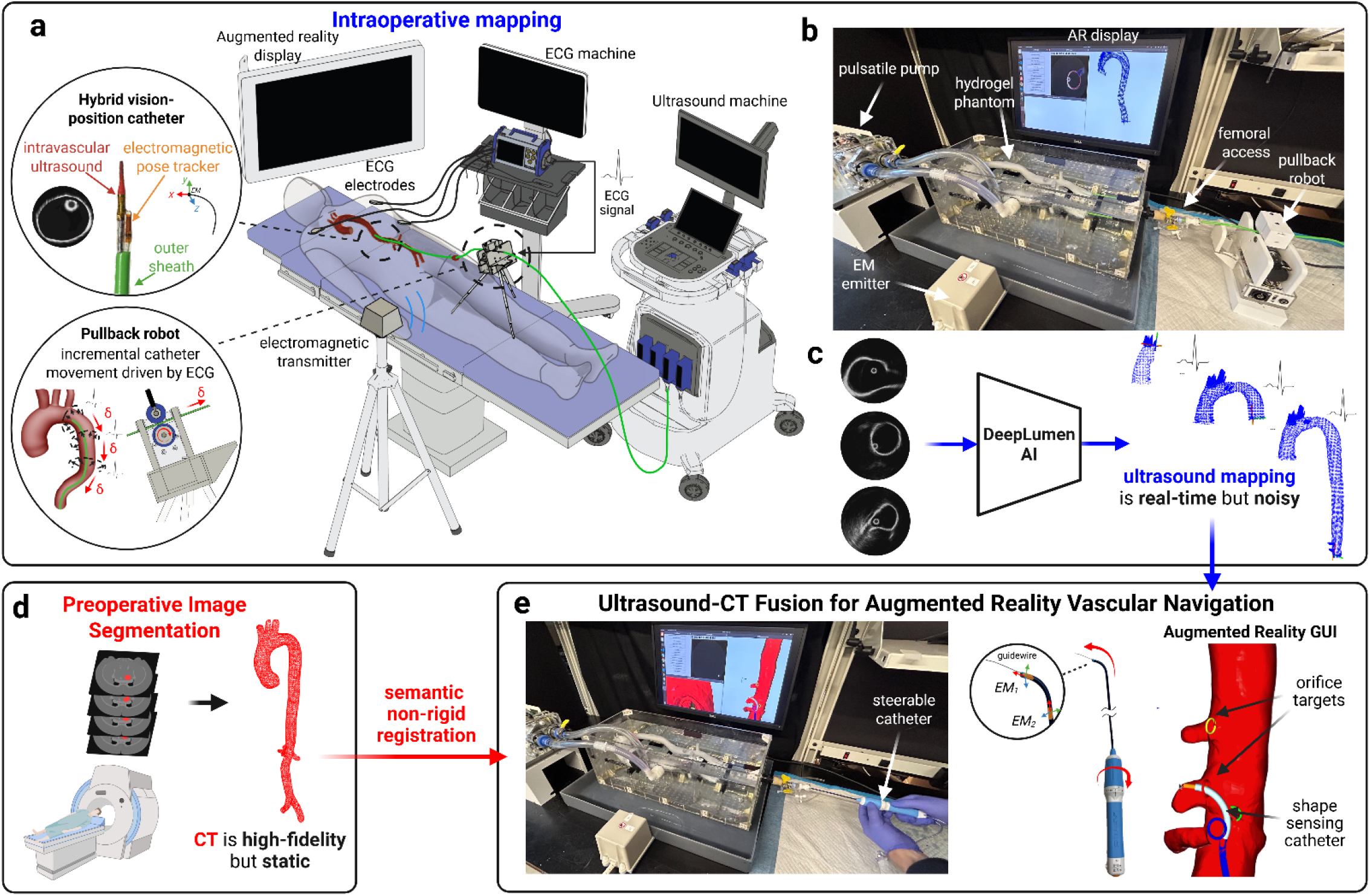
Platform for augmented reality (AR) endovascular surgery driven by multi-modal fusion of ultrasound, position sensing, and preoperative data. **a**. An intravascular ultrasound (IVUS) catheter is coupled to an electromagnetic (EM) position sensor to yield a hybrid catheter with both vision and position sensing capabilities suitable for vascular mapping. The catheter is retracted through the vasculature by a robotic device that synchronizes catheter motion with the patient’s heartbeat, as measured by the electrocardiogram (ECG) signal. **b**. The clinical setup is replicated in vitro using patient-specific aortic phantom models driven by pulsatile flow. **c**. EM measurements are combined with ultrasound images segmented by a deep learning algorithm to obtain a 3D or 4D map of the vasculature. **d**. The intraoperative reconstruction of the vasculature is fused with preoperative computed tomography (CT) imaging via non-rigid registration to yield a higher-fidelity reconstruction suitable for real-time AR procedural guidance. **e**. Electromagnetic sensors are integrated with an off-the-shelf steerable catheter sheath, enabling real-time localization of the sheath tip within the AR user interface and, in turn, guidance for endovascular procedures such as target vessel cannulation (TVC). The graphical UI displays: the global 3D view of the reconstructed aorta (red), catheter shaft (dark blue rigid segment, light blue steerable segment), and branch ostia (colored rings). Supplementary Video S1 provides a visual overview of the AR platform and its operation.

Our work makes several advances. We introduce DeepLumen, a deep learning architecture for IVUS segmentation pretrained on patient-specific phantoms and fine-tuned for large-animal imaging, as well as a semantically driven non-rigid registration framework that corrects deformation between noisy IVUS-EM reconstructions and high-fidelity CT. We extend this framework to cardiac-driven motion capture using ECG gating and time-synchronized robotic pullback. We validate the platform in vitro and in vivo, benchmarking AR TVC against fluoroscopic guidance, and demonstrate its application to a fenestrated EVAR. To our knowledge, no prior work has applied IVUS-EM reconstructions to clinically meaningful tasks such as TVC or EVAR in preclinical settings. We release our in vitro and in vivo 3D datasets (https://doi.org/10.5281/zenodo.20737791) to support the development of registration algorithms for IVUS-based navigation. Moreover, we release labeled IVUS imaging (https://doi.org/10.5281/zenodo.20752361) of the aortic lumen and branch for researchers developing AI algorithms for IVUS.

## Results

### DeepLumen Segmentation Framework

Classical IVUS lumen and branch segmentation methods, such as spline-based boundary fitting^20,22^ and peak detection^27^, are vulnerable to imaging artifacts. We developed DeepLumen, a convolutional neural network (CNN) architecture with an augmented receptive field to model long-range noise artifacts typical of IVUS^28^ (e.g., reverberation, acoustic shadowing), while preserving the inductive biases that make CNNs robust on small datasets (architectural details in Methods). To overcome a lack of publicly available IVUS data (the largest dataset contains 326 annotated frames^29^) and reduce reliance on costly animal data, the model was pretrained on IVUS images from ultrasound-compatible, patient-specific aortic phantoms (Fig. 2a). Unlike previously reported idealized hydrogel aortic phantoms^30^, our fabrication method captures the anatomical complexity, branch configurations, and tortuosity of patient-specific aortic anatomy (see Methods). We conducted ablation studies across three U-Net variants (Fig. 2b), using standard U-Net as the baseline, adding mechanisms for increased receptive field and spatial attention while maintaining CNN inductive biases. Fig. 2c shows Dice coefficient (DSC) performance for lumen and branch segmentation across all four architectures (n=7 phantoms). No significant difference in lumen segmentation was observed as expected, given it is easily identifiable and occupies a large image footprint. In contrast, branch segmentation performance varied significantly across architectures (one-way ANOVA, F(3,24) = 7.28, p = 0.0012), with DeepLumen achieving the highest DSC (baseline: 0.640±0.068, ViT-UNet: 0.712±0.083, DRN-UNet: 0.739±0.059, DeepLumen: 0.805±0.052). DeepLumen significantly outperformed the baseline (p = 0.0006) and DRN-UNet (p = 0.049), while the improvement over ViT-UNet approached but did not reach significance (p = 0.069). All modified architectures substantially exceed the empirically-determined DSC threshold (>0.6) sufficient for robust non-rigid registration downstream in our pipeline. We validated the DeepLumen architecture using preclinical large animal IVUS imaging (n=7 animals), to determine if DeepLumen pretrained on IVUS images from patient-specific aortic phantoms improves segmentation performance in vivo, despite visual differences between ultrasound-compatible phantoms and native aortic tissue (Fig. 2d). Three training strategies were compared: training from scratch, joint training on combined phantom and animal data, and phantom pretraining with animal fine-tuning (Fig. 2d). Branch segmentation performance varied significantly among strategies (one-way ANOVA, F(2,15) = 6.20, p = 0.008), where fine tuning achieved the highest DSC (0.654 ± 0.0366), followed by joint training (0.608 ± 0.084) and training from scratch (0.536 ± 0.067).

**Fig. 2.**
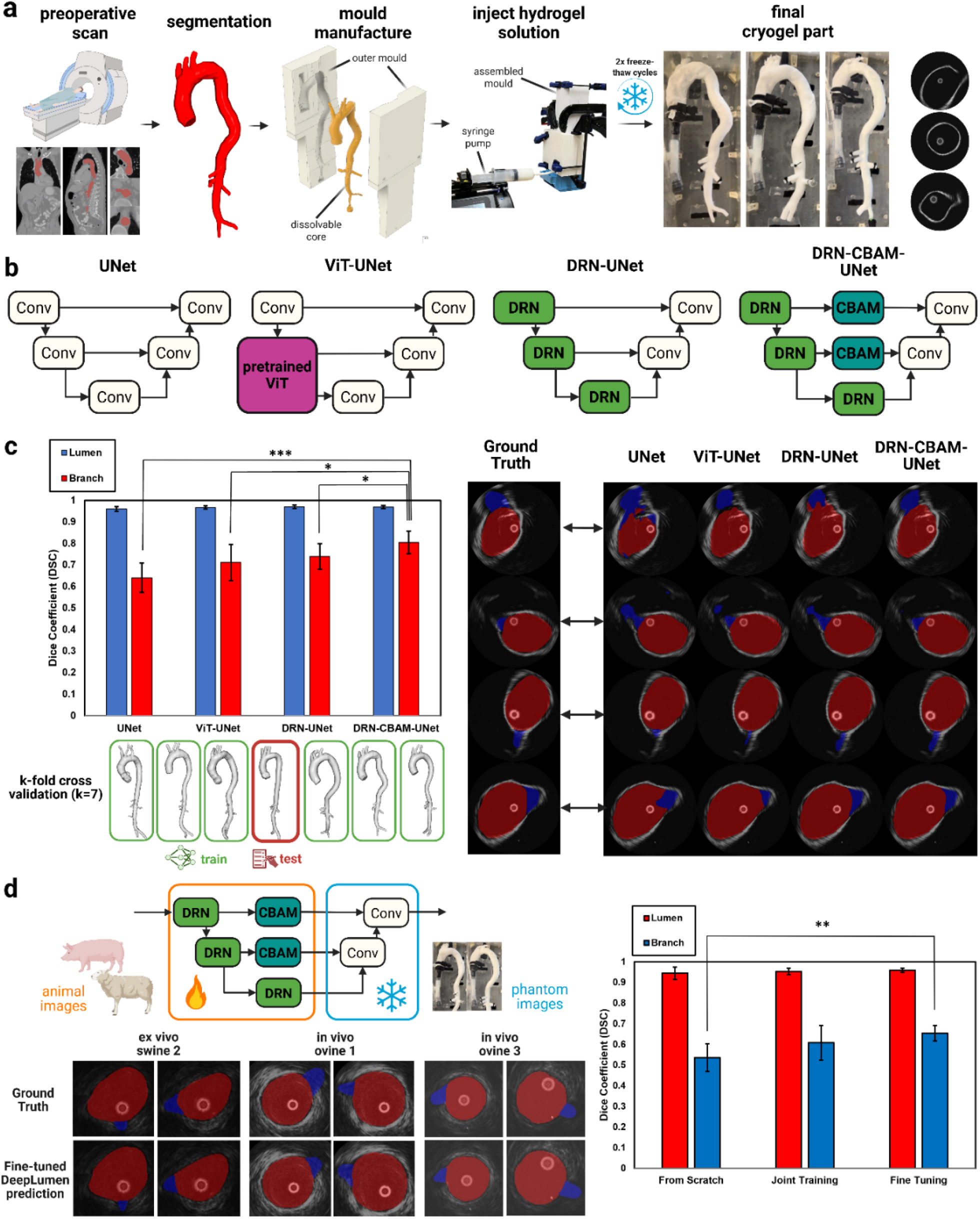
DeepLumen IVUS segmentation. **a**. Patient-specific hydrogel aortic phantom fabrication pipeline, consisting of CT segmentation, mould design, hydrogel injection moulding, and freeze–thaw cycles. **b**. Four deep learning architectures (simplified for illustrative purposes) were evaluated for IVUS segmentation: a standard U-Net baseline; a hybrid CNN–Transformer (ViT-UNet) with a pretrained ViT encoder; a Dilated Residual U-Net (DRN-UNet); and an Attention-Enhanced U-Net (DRN-CBAM-UNet) combining dilated convolutions with a convolutional block attention module. **c**. Quantitative and qualitative comparison of segmentation performance across network architectures in vitro. Dice coefficient (DSC) scores for lumen and branch segmentation were evaluated using k-fold cross-validation (k = 7, n = 7 patients). Expert-annotated ground truth labels and corresponding network predictions are shown. **d**. Quantitative and qualitative large-animal IVUS segmentation performance across three training strategies: training from scratch, joint training, and fine-tuning. For fine-tuning, a transfer learning strategy was employed in which the decoder layers of the DeepLumen architecture are frozen to facilitate fine-tuning of the encoder towards animal imaging. Error bars denote ± 1 standard deviation (SD). * denotes statistical significance ( p < 0.05, ** p < 0.005; Tukey’s post hoc test).

### IVUS-CT Non-Rigid Registration Framework

3D IVUS-EM reconstructions, obtained using signed distance function (SDF)^35,36^ mapping (detailed in Methods), produce geometrically plausible aortic surfaces but remain noisy and may include false branch detections. We developed a semantic non-rigid registration algorithm for CT-IVUS fusion that achieves automated global alignment without fiducial markers or manual initialization, despite non-rigid deformation and false branch detections. Our pipeline (described in Methods) comprises three sequential stages:

*(1)rigid registration* with Iterative Closest Point (ICP)^39^ for coarse global alignment between the two geometries; (2) *correspondence estimation* with a Hidden Markov Model (HMM)^40^ that leverages semantic structural information while rejecting outliers; (3) we *deform the preoperative mesh* onto the intraoperative IVUS geometry using a variant of non-rigid ICP^41^. Fig. 3 illustrates representative results for two in vitro registrations. Orifice deviation between CT and IVUS branch centroids was computed for the 4 major abdominal branches where TVC is typically performed. We conducted ablation studies to compare our HMM correspondence estimation technique against NN and Random Sample Consensus (RANSAC). We perform 50 iterations of RANSAC, requiring approximately 5 minutes of computation time, which is comparable to the duration for preoperative-to-intraoperative image registration^42^. For each patient, 3 independent IVUS pullbacks were acquired and registered to the preoperative CT. A one-way ANOVA revealed a significant effect of correspondence method on orifice deviation (F(2,233) = 47.87, p < 0.0001). HMM-based correspondence achieved significantly lower orifice deviation than both NN (1.66±1.07 mm vs. 11.44±6.53 mm, p < 0.0001; N = 4 branches × 3 pullbacks × 7 phantoms) and RANSAC (9.46±9.33 mm, p < 0.0001), with most branch registrations achieving deviation within 1-2 mm; well below the previously mentioned clinically-accepted threshold of 3 mm for accurate device positioning during TVC.

**Fig. 3.**
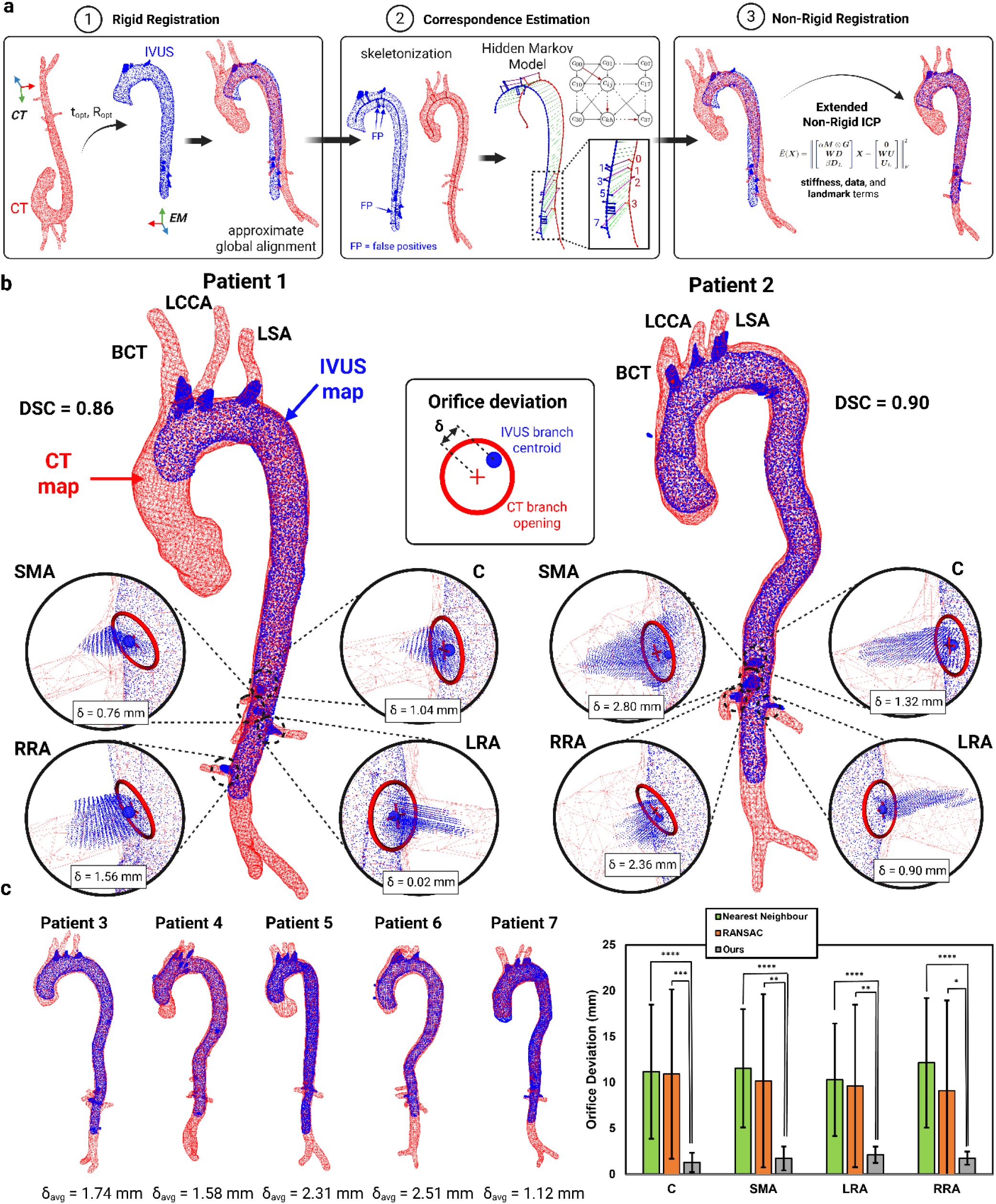
CT–IVUS non-rigid registration framework. **a**. Non-rigid registration pipeline, comprising rigid alignment of CT with the IVUS reconstruction using iterative closest point (ICP), correspondence estimation via a Hidden Markov Model (HMM), and non-rigid deformation of the CT geometry onto the IVUS– EM reconstruction using an extended non-rigid ICP formulation. **b**. Representative registration results for Patient 1 and Patient 2, showing the raw IVUS reconstruction (blue) and registered CT surface (red). Orifice deviation (δ) quantifies the Euclidean distance between the CT branch opening centroid (red crosshairs) and its corresponding IVUS branch centroid (blue sphere). **c**. Quantitative registration performance across all patient cases. Orifice deviation for each of the four major abdominal aortic branches across seven patients, comparing three correspondence estimation methods: nearest neighbour (NN), RANSAC, and the proposed HMM-based method (Ours). Error bars denote ± 1 standard deviation (SD). *, **, ***, **** denote statistical significance (Tukey’s post hoc test). Branch abbreviations: SMA, superior mesenteric artery; C, celiac trunk; LRA, left renal artery; RRA, right renal artery; BCT, brachiocephalic trunk; LCCA, left common carotid artery; LSA, left subclavian artery.

### ECG-gated motion capture pipeline

To capture cardiac-driven aortic deformation which may affect TVC ease, we extended our framework to 4D using ECG-gated robotic pullback. We developed an automated pullback robot that synchronizes catheter translation and facilitates both image and EM sensor data gating based on the patient’s ECG signal. ECG-gated robotic pullback bins IVUS frames and EM poses by cardiac phase, producing multiple phase-specific 3D reconstructions (Fig. 4), to which our non-rigid registration pipeline is applied sequentially. The CT is first registered to the end-diastolic IVUS reconstruction (phase 0), with the resulting deformed mesh serving as initialization for registration to phase 1, which is then propagated through all subsequent phases to capture the full 4D motion field of the aorta. Example outputs are shown for 3 representative dynamic *in vitro* phantoms with pulsatile flow applied in Fig. 5. To quantitatively validate our 4D motion tracking accuracy, we compared IVUS-derived motion trajectories against ground truth data acquired using a clinical 4D-CT scanner for ECG-gated contrast scans of 3 *in vitro* phantoms with pulsatile flow. Time-averaged errors across all cardiac phases were: Patient 2: 1.36±1.0 mm, Patient 4: 1.59±1.32 mm, Patient 6: 1.88±1.48 mm. Videos showing sample outputs from the motion capture pipeline and our corresponding validation protocol are shown in Supplementary Video S3. The clinical impact of motion compensation on TVC performance is addressed in the preclinical validation section.

**Fig. 4.**
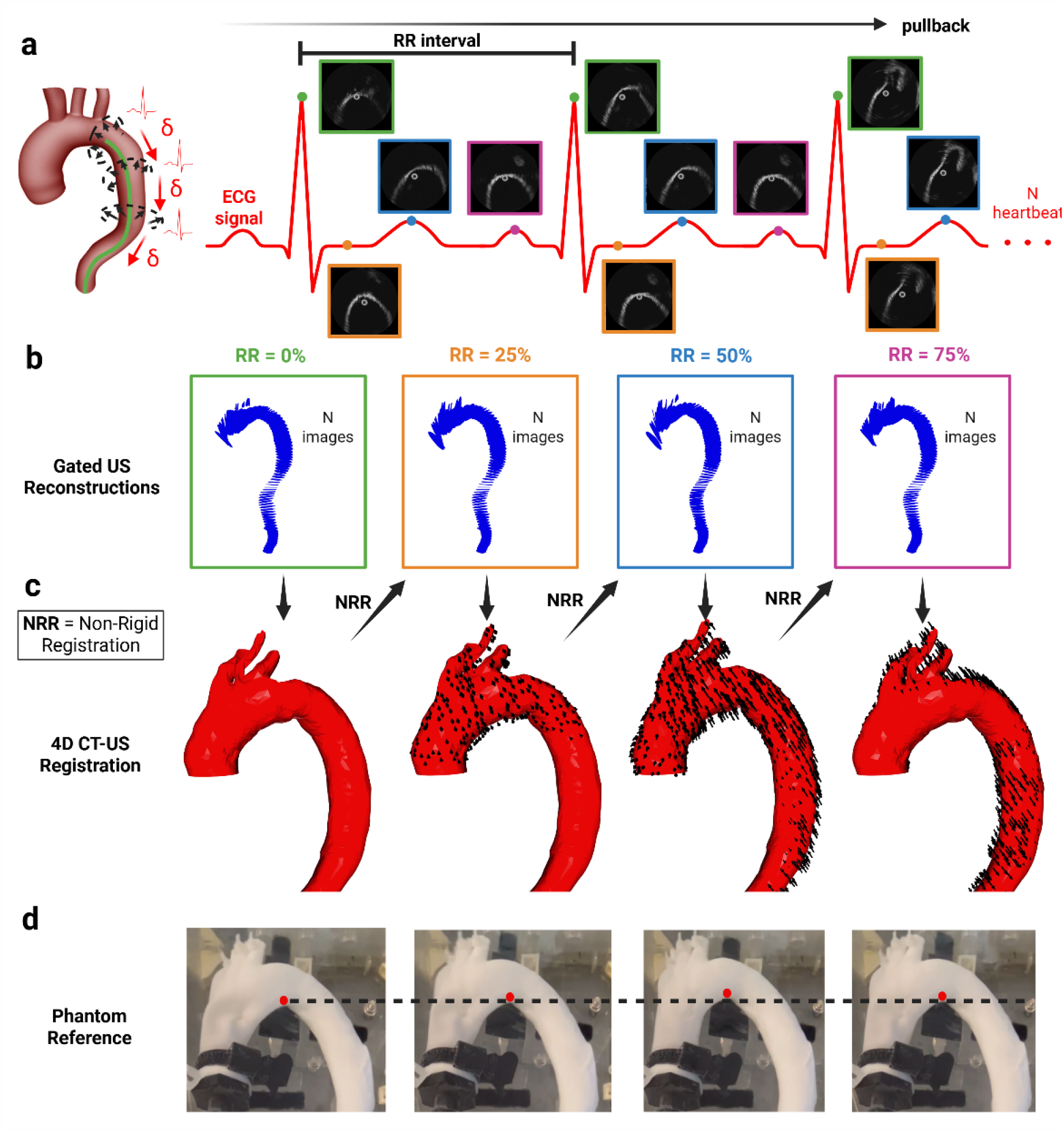
ECG-gated motion capture pipeline. **a**. The automated pullback robot synchronizes catheter translation with the ECG signal, enabling IVUS frames and EM poses to be binned by cardiac phase according to their acquisition timing within the cardiac cycle. **b**. Each phase-specific bin yields a 3D aortic reconstruction exhibiting noise and spatial discontinuities due to limited sampling density and ultrasound artifacts. **c**. The preoperative CT is first registered to the end-diastolic phase using the non-rigid registration pipeline described in Fig. 3a, then sequentially to each subsequent phase, maintaining geometric smoothness while accurately tracking aortic wall motion. Motion vectors (arrows) illustrate vertex displacement trajectories throughout the cardiac cycle. Readers are strongly encouraged to view Supplementary Video S3, which provides an animated visualization of the motion capture framework illustrated in this plot. **d**. A tracked point on the aortic arch (red marker) illustrates the motion pattern of the patient-specific hydrogel phantom for reference.

**Fig. 5.**
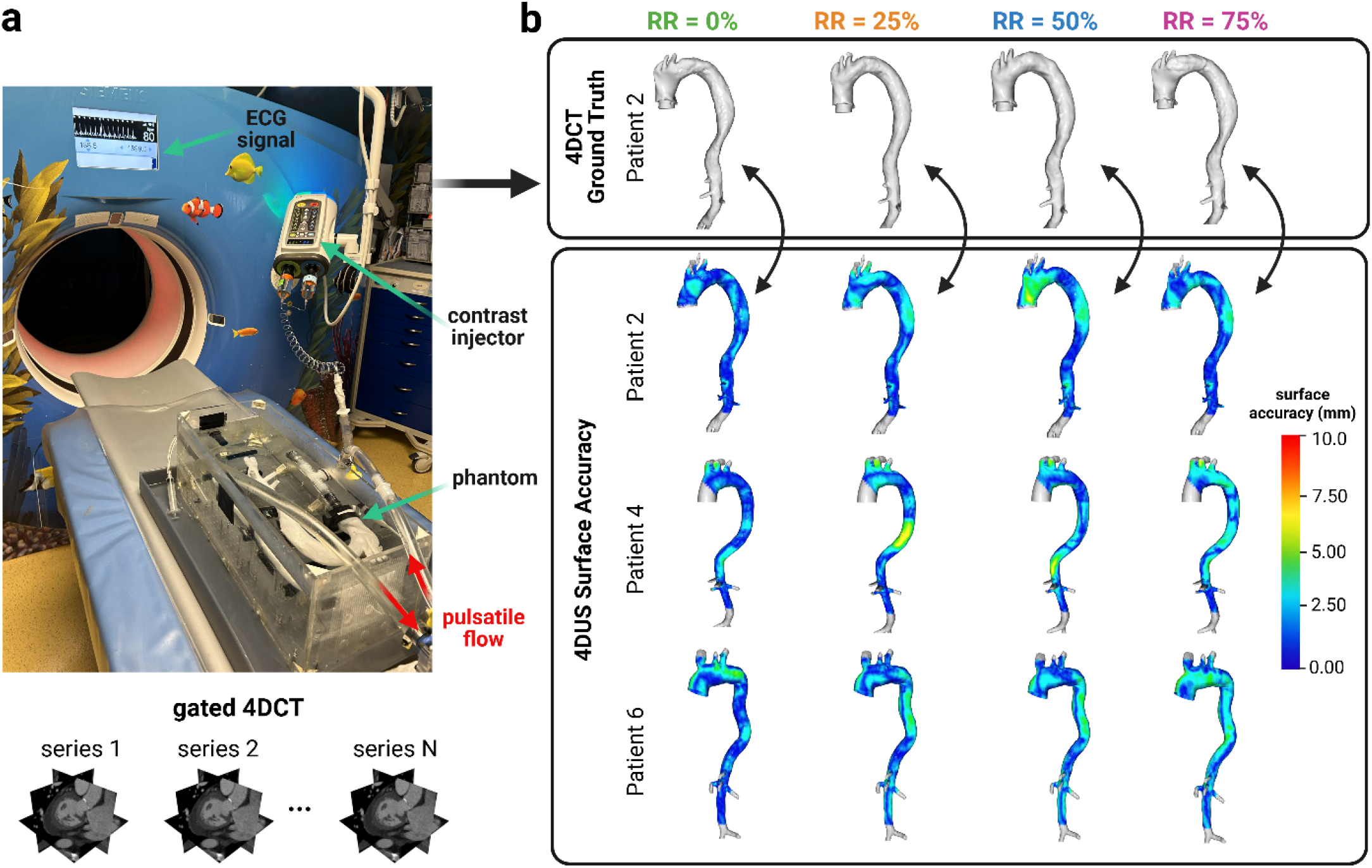
Quantitative validation of 4D-US registration against 4D-CT. **a**. Apparatus for ECG-gated contrast-enhanced 4DCT scanning of an in vitro hydrogel phantom. The output consists of multiple 3D reconstructions of the phantom, each representing a distinct phase of the cardiac cycle. **b**. Segmentations derived from the 4DCT scan are compared to the 4D-US registration, and a bidirectional surface distance map is computed for each patient at each cardiac phase. Results at RR = 0%, 25%, 50%, and 75% are shown for illustrative purposes. Readers are strongly encouraged to view Supplementary Video S3, which provides an animated visualization of the surface-contour results shown above.

### Preclinical validation studies

We instrumented a commercial steerable sheath (TourGuide, Aptus EndoSystems) with two EM sensors (Aurora, NDI) and a guidewire delivery channel, integrated with a real-time AR interface built on Open3D^43^ and ROS^44^. Four vascular surgery residents (post-graduate years 3-5) performed TVC under two conditions: (1) standard fluoroscopic guidance and (2) AR guidance, with fluoroscopy restricted to a brief confirmatory shot after each cannulation. Guidance order was counterbalanced across phantoms to minimize learning effects. As described in the following sections, performance was assessed across a range of preclinical contexts for four metrics: cannulation time, cumulative fluoroscopy exposure, number of attempts per vessel (a proxy for trauma risk), and NASA Task Load Index^45^ for cognitive demand.

#### 1. in vitro study

Fig. 6 summarizes the quantitative results of the 7 *in vitro* phantom experiments. Overall *cannulation times* were significantly shorter with AR guidance compared to fluoroscopy (AR: 21.4±14.5s, X-ray: 32.9±22.3s; paired t-test, p = 0.0017, N = 112 cannulations), *fluoroscopy exposure* was dramatically reduced with AR guidance (AR: 3.2±1.6s, X-ray: 31.7±22.0s; p < 0.001, N = 112 cannulations), as were *cannulation attempts* (AR: 1.27±0.54 attempts, X-ray: 2.64±2.49 attempts; p < 0.001, N = 112 cannulations) and *NASA Task Load Index* for all metrics related to cognitive demand (see Supplementary Text S3 for details). The dual-view interface enables a two-stage alignment strategy, whereby the surgeon first orients the catheter toward the target branch (visualized as a colored ring) using the global perspective, then switches to the endoscopic view for precise tip-to-orifice alignment (Fig. 6c). Furthermore, we analyzed learning curves for cannulation time across the seven sequential phantoms for each surgeon (see Supplementary Text S4). Finally, to quantify the benefit of dynamic motion tracking over static registration, we compared cannulation performance using 4D motion-compensated AR guidance versus conventional 3D static guidance (see Supplementary Text S5). No significant difference between 4D motion-compensated guidance and 3D static guidance was observed, suggesting that orifice motion does not affect cannulation for abdominal TVC.

**Fig. 6.**
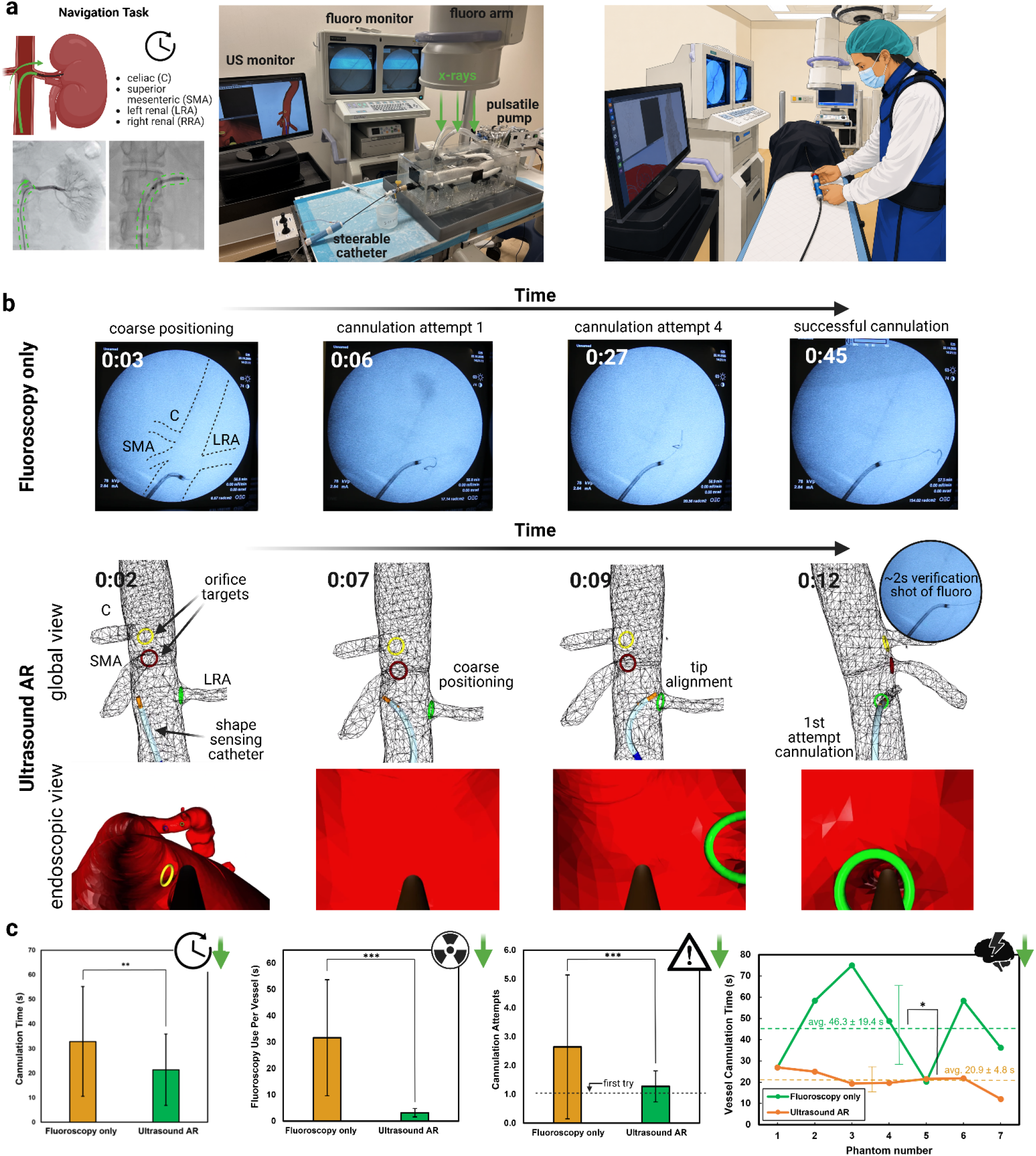
In vitro augmented reality (AR) platform preclinical validation. **a**. Study design for comparative evaluation of the AR platform against fluoroscopy-only guidance. Four vascular surgery residents cannulated four major abdominal branches (celiac, SMA, left renal, and right renal arteries) across seven patient-specific phantoms under both X-ray and AR guidance conditions (N = 112 total cannulations per condition), with performance assessed via cannulation time, radiation exposure, number of attempts, and cognitive workload. **b**. Qualitative comparison of cannulation workflows under fluoroscopic and AR guidance. Fluoroscopic guidance requires continuous radiation exposure as the surgeon makes repeated trial-and-error attempts to cannulate branches visualised only as 2D projections. AR guidance provides continuous 3D visualisation of catheter position relative to branch orifice targets (coloured rings). **c**. Quantitative comparison of target vessel cannulation performance under fluoroscopic and AR guidance. Cannulation time, fluoroscopy exposure duration, and number of cannulation attempts per branch are shown. Cannulation time trajectories across the seven in vitro phantoms illustrate performance evolution with increasing operator experience, averaged across all four surgical residents. Dashed lines indicate mean performance across all phantoms. Error bars denote ± 1 standard deviation. *, **, *** denote statistical significance (Tukey’s post hoc test). Supplementary Video S5 provides a visual demonstration of the X-ray and AR interfaces.

#### 2. in vivo study

To establish clinical translatability, we performed the first IVUS-guided TVC experiments in a living animal model across three ovine subjects. A preoperative CT scan of the sheep was obtained ∼48 hours before the procedure (procedural details in Methods). Mean orifice deviation between CT and IVUS branch centroids (Fig. 7c) was 1.1±0.9 mm for Sheep 1, 1.88±0.4 mm for Sheep 2, and 1.53±0.73 mm for Sheep 3; well below the previously mentioned 3 mm clinical tolerance threshold established for accurate device positioning. Two operators (a PGY-4 vascular surgery resident and an electrophysiologist with extensive catheter-based experience) performed TVC of all four abdominal branches under both fluoroscopic and AR guidance conditions (N = 24 cannulations per condition: 4 branches × 2 operators × 3 sheep). Mean cannulation time was significantly reduced with AR guidance across the experiments (AR: 30.2±19.7s, X-ray: 75.8±76.0s; paired t-test, p = 0.0083). Fluoroscopic use was also significantly reduced (AR: 4.0±2.71s, X-ray: 64.3±64.6s; p = 0.0013), and the number of cannulation attempts differed significantly between conditions (AR: 1.42±0.64, X-ray: 2.95±1.75; p = 0.0019).

**Fig. 7.**
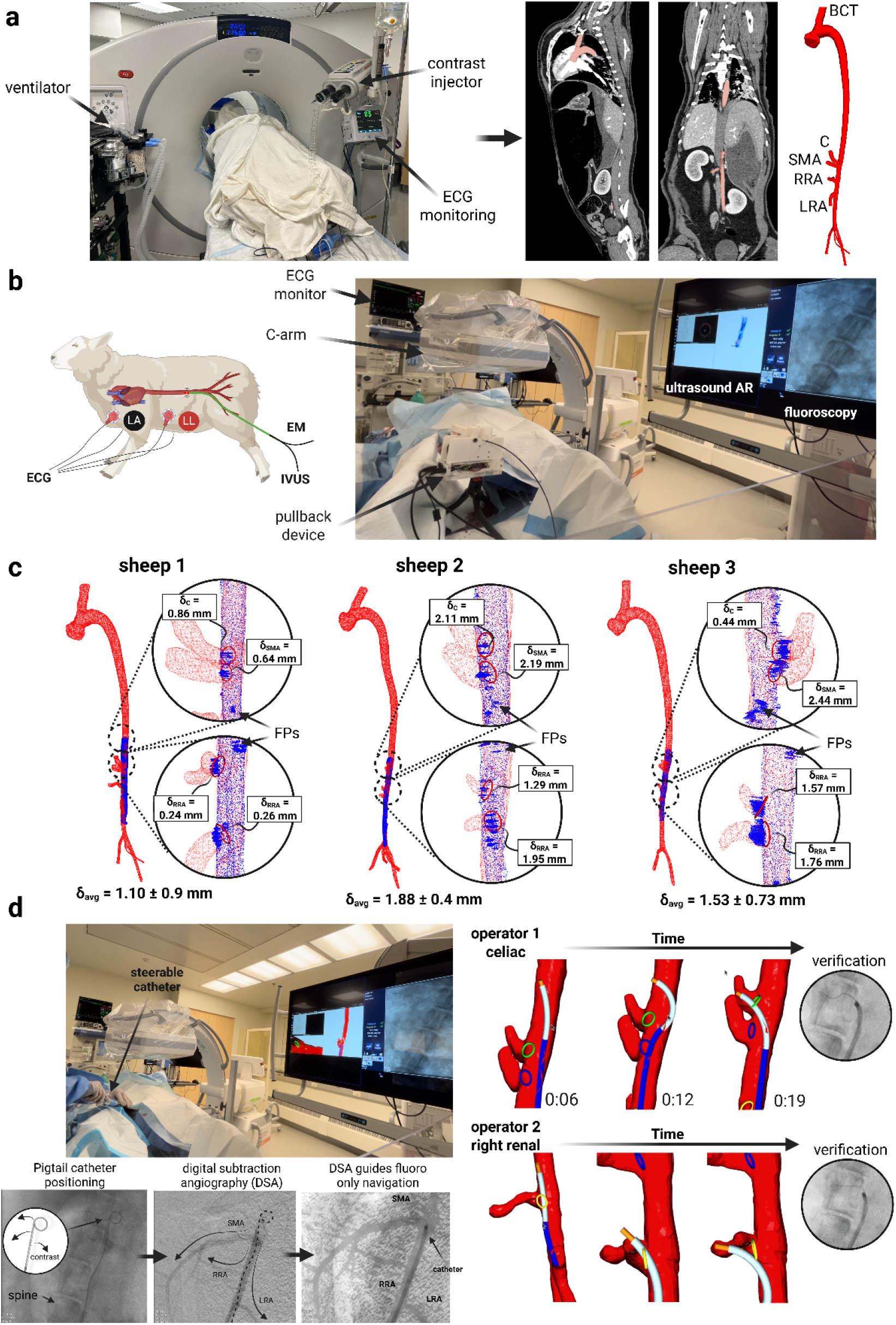
In vivo augmented reality (AR) platform preclinical validation. **a**. ECG-gated contrast-enhanced preoperative CT imaging of an ovine model. The aorta and major branch vessels are segmented from the CT angiogram prior to the interventional procedure. **b**. Interventional suite configuration for in vivo ovine studies. The fluoroscopy C-arm is positioned over the animal’s abdomen to visualise the major abdominal branch vessels. Four-lead ECG monitoring (RA: right arm, LA: left arm, RL: right leg, LL: left leg) provides cardiac synchronisation for automated ECG-gated IVUS pullback. **c**. CT–IVUS registration results for all three animals, showing the CT and IVUS geometries and corresponding orifice deviations (δ). **d**. In vivo ovine validation of ultrasound-guided AR navigation. For fluoroscopic guidance, digital subtraction angiography (DSA) was performed by positioning a 5 Fr pigtail catheter in the descending thoracic aorta to deliver iodinated contrast into the abdominal aorta. The resulting DSA roadmap was overlaid as a semi-transparent mask on live fluoroscopy to provide anatomical reference for target vessel cannulation. Representative AR-guided cannulation sequences for the celiac and right renal arteries are shown. Branch abbreviations: BCT, brachiocephalic trunk; C, celiac; SMA, superior mesenteric artery; LRA, left renal artery; RRA, right renal artery. In accordance with Institutional Animal Care and Use Committee (IACUC) guidelines on research animal photography, the animal was draped during photography. Supplementary Video S7 provides a visual demonstration of the preoperative imaging and surgical procedure conducted during in vivo experiments.

### FEVAR demonstration

Fenestrated endovascular repair (FEVAR) extends standard EVAR to aneurysms involving major abdominal branches^46^, requiring the operator to navigate through fenestrations (i.e., openings) in the deployed graft before cannulating each branch vessel. During FEVAR, TVC is the most radiation-intensive phase of the procedure^6,47^. As a capstone application of our platform, we demonstrate an AR workflow integrable with physician-modified endograft planning (PMEG)^48^. We affixed highly echogenic tungsten beads (0.8 mm diameter) adjacent to each fenestration, which serve as fiducial markers detectable via IVUS (see Methods for integration details). By identifying bead positions with IVUS, we reconstruct the deployed graft geometry including fenestration locations in 3D and a virtual model of the graft can be superimposed onto the registered aneurysm anatomy for 3D AR guidance. Four vascular surgery residents performed target vessel cannulation with the fenestrated graft under both fluoroscopic and AR guidance conditions, attempting all four branch vessels (N = 16 total cannulations per condition). Mean cannulation time was reduced by 30% (AR: 99.1±68.7 s, X-ray: 143.2±73.6 s; paired t-test, p = 0.138) and mean cannulation attempts per branch by 38% (AR: 2.56±1.79, X-ray: 4.0±1.63; p = 0.065), both approaching but not reaching significance. Notably, AR guidance achieved a significant reduction in fluoroscopy exposure (AR: 6.7±6.2 s, X-ray: 113.5±61.9 s; p < 0.001).

## Discussion

Multimodal fusion of IVUS, EM tracking, and preoperative CT enables real-time AR guidance that demonstrably reduces operative time, radiation exposure, vessel trauma, and cognitive burden during endovascular navigation. We proposed a deep learning framework, DeepLumen, for achieving robust in vivo IVUS image segmentation despite limited animal datasets. We hypothesize that DeepLumen’s advantage over the ViT-based variant stems from preserving the inductive biases of CNNs; whereas ViT’s scaled dot-product attention tends to overfit on small biomedical datasets, DeepLumen extends the receptive field for long-range artifacts while retaining the convolutional priors that generalize from limited data. This aligns with reports that task-specific U-Nets can outperform large vision-transformer models such as MedSAM on specialized medical segmentation tasks, where training data is scarce^49^. Phantom pretraining with animal fine-tuning improved in vivo branch segmentation over training from scratch, reducing reliance on costly animal datasets. Branch segmentation was shown to be more challenging in vivo than in vitro due to artifacts from adjacent anatomical structures compared to a controlled phantom environment. Our fully supervised training paradigm still currently requires manual annotation of IVUS images, which limits dataset scale. Automated synthetic IVUS generation from CT volumes could scale labeled training data without additional annotation effort^50^.

We presented the first automated non-rigid CT-IVUS registration framework capable of fusing preoperative and intraoperative anatomy without fiducial markers or manual initialization. The HMM-based correspondence estimation significantly outperformed nearest-neighbor matching and RANSAC. In comparison to random sampling with RANSAC, HMM efficiently identifies anatomically plausible correspondence sequences by encoding semantic constraints that prune the search space and converge to the global optimum. Moreover, we presented the first 4D motion capture framework driven by 2D ultrasound with position tracking with temporal tracking errors in the 1-2 mm range across the cardiac cycle, well below the previously cited 3 mm heuristic used by clinicians to facilitate TVC, as well as the 4–10 mm error range reported for intraoperative CT-to-fluoroscopy registration methods. While evaluation of 4D motion compensation demonstrated that cardiac-driven motion tracking provides minimal benefit in the abdominal aorta, high-motion applications such as aortic valve repair and thoracic EVAR may benefit.

We validated these technical capabilities through comprehensive preclinical testing, demonstrating improvements across all clinical metrics. X-ray lacks depth cues, requiring extensive trial-and-error to align the catheter tip with branch ostia. The AR system also represents a drastic reduction in radiation dose per cannulation as expected, given AR restricts its use to brief confirmatory shots rather than continuous exposure throughout navigation. EM tracking was selected over FOSS due to its greater accessibility in research settings. FOSS integration could further eliminate the need for fluoroscopic verification altogether, as it enables direct measurement of guidewire shape and position within branch vessels. Rapid skill acquisition is particularly encouraging given all participants had 3+ years of prior fluoroscopy experience, suggesting the AR interface is highly learnable despite its novelty. We further demonstrated generalizability of the non-rigid registration framework to living tissue, achieving a drastic reduction in fluoroscopy. Notably, in Sheep 1, contrast injection (Omnipaque 350 mgI/mL, 30 mL bolus) induced anaphylactic shock, which resolved within 5 minutes after epinephrine administration (0.3 mg intramuscularly). In Sheep 3, an abdominal aortic dissection occurred during attempted fluoroscopy-guided TVC, preventing LRA cannulation and requiring exclusion of the LRA from analysis. Together, these adverse events underscore the motivation for contrast-free navigation platforms that improve three-dimensional spatial awareness during TVC, particularly in patients with renal insufficiency or prior contrast allergy. Finally, to emphasize clinical applicability, we demonstrated the first use of IVUS for 3D navigational guidance for fenestrated endovascular aneurysm repair and observed dramatic reductions in fluoroscopy use, illustrating how the platform slots into one of the most demanding real-world procedures. Given the substantial cost of fenestrated devices (∼$18,000 per graft^51^), this study was designed as a proof-of-concept evaluation using a single device rather than preclinical validation across multiple patient-specific grafts. Nevertheless, the observed trends were directionally consistent with the larger in vitro and in vivo studies, supporting the potential of AR guidance to improve FEVAR navigation.

## Outlook

By leveraging existing or readily deployable modalities such as IVUS and CT, the AR platform may reduce capital and infrastructure barriers to clinical translation, improving the accessibility of complex interventions such as FEVAR. Beyond aortic intervention, the 4D framework generalizes to any endoluminal procedure with periodic motion and an available gating signal, such as navigational bronchoscopy gated by spirometry, contributing to a broader vision of real-time digital twins for patient-specific procedural guidance. These capabilities may also be relevant to future robotic systems: Dupont et al. identify “time-critical endoluminal interventions,” such as TVC, as “low-hanging fruit” for first-generation autonomous surgical robots^52^. By augmenting three-dimensional spatial awareness beyond conventional X-ray guidance, our AR platform could support the real-time perception-planning-action loop required for conditional autonomy, thereby extending the value proposition of endovascular robotics beyond prior manually controlled systems such as Magellan (Hansen Medical, CA, USA).

## Methods

### System hardware components and inter-process communication (IPC)

A Lenovo ThinkPad X1 laptop equipped with an Intel Core i9 processor, 64 GB RAM, and NVIDIA GeForce RTX A5500 GPU running Ubuntu Linux 20.04 was used for the study. We use the Robot Operating System (ROS) as the middleware framework^44^, enabling parallelized sensor polling, motor actuation, real-time mapping, and inter-node communication. Visualization is performed using the Open3D library^43^. IVUS imaging is performed using a Volcano S5 imaging console (Philips, NV, Netherlands). For *in vitro* benchtop experiments, we use a Philips 0.035” PV Visions 10 MHz IVUS catheter, while *in vivo* animal studies utilize a Philips 0.018” PV Visions 30 MHz catheter to accommodate smaller vessel diameters. IVUS images are streamed from the IVUS machine to the computer using an Epiphan AV.io HD video capture card (Epiphan, Ontario, CA). 6DOF EM tracking is provided by an Ascension trakSTAR electromagnetic tracking system with model 180 sensors (2 mm diameter). Cardiac ECG gating is achieved using a Dash 4000 patient monitor (GE Healthcare). For *in vitro* benchtop experiments where no physiological signal is available, the Dash monitor is replaced with a HeartRoid pulsatile flow pump, which generates a synthetic ECG-like trigger signal correlated with the pump’s cyclic motion. Automated catheter pullback coordination with the ECG signal is controlled by a custom Arduino-based microcontroller embedded in the pullback robot. A diagram showing the entire IPC framework is shown in Supplementary Figure S4.

### Patient-specific hydrogel aortic model manufacture pipeline

Patient-specific aortic phantoms were fabricated from polyvinyl alcohol (PVA) hydrogel, selected for its tissue-matched acoustic and mechanical properties (Supplementary Text 8), using the following workflow, illustrated in Fig. 2a:

#### (1) Geometric segmentation

The aortic lumen is segmented from preoperative CTA data using semi-automated tools in 3D Slicer^60^.

#### (2) Mold design and fabrication

A multi-part outer mold and a water-soluble inner core (representing the negative space of the vessel lumen and branches) are designed in CAD software. Exact-constraint kinematic coupling principles^61^ are incorporated into the mold design to ensure precise alignment between inner core and outer mold halves, preventing deflection that would compromise surface quality. Both components are additively manufactured using a fused deposition modeling (FDM) printer (Bambu Lab P1S, Bambu Studio). The outer mold is printed in polylactic acid (PLA) filament and the inner core in water-soluble PVA filament.

#### (3)Hydrogel preparation

A PVA hydrogel solution is prepared with a mass ratio of 20:5:100 for PVA powder, calcium carbonate powder, and deionized water, respectively. The solution volume is calculated based on the void volume between the assembled core and outer mold. The mixture is continuously heated to boiling with constant stirring to fully dissolve the PVA powder and prevent agglomeration. Calcium carbonate is added as an acoustic scattering agent to increase ultrasound backscatter and better simulate tissue echogenicity^30^.

#### (4)Injection molding

After assembling the water-soluble core within the multi-part outer mold, the heated PVA solution is injected under pressure through inlet ports using a syringe pump. Riser channels at the mold apex allow air evacuation and provide visual confirmation of complete filling.

#### (5)Freeze-thaw cycling

Two complete freeze-thaw cycles (24 hours each) are performed to enhance crystallinity and tune the mechanical stiffness of the hydrogel. Each cycle consists of 12 hours at -20°C followed by 12 hours at room temperature.

#### (6)Core dissolution

After the second thaw, the outer mold is disassembled and the PVA inner core is dissolved by placing the vessel and core in a water bath for a further 12 hours.

### Data collection and labeling

IVUS imaging data was acquired from seven patient-specific aortic phantoms fabricated from geometries in the Aortic Vessel Tree (AVT) CTA dataset^62^, an open-source anonymized multicenter database containing contrasted aortic scans from diverse geographic locations that captures the morphological variability of human aortic anatomy. Each phantom underwent a full-length pullback acquisition using a Philips Visions PV 0.035” IVUS catheter imaging at 30 Hz (600 images per phantom) yielding approximately 4,400 raw images in total. Images were manually annotated using the Labelbox collaborative annotation platform (Labelbox Inc, San Francisco, CA) by a team of two biomedical engineers with expertise in ultrasound interpretation and one cardiothoracic surgical resident. Manual labeling by domain experts was chosen to embed clinical knowledge about boundary identification in the presence of acoustic artifacts. The semantic classes labeled were the vessel lumen (defined as the largest approximate elliptical region enclosed by the intimal boundary visible in IVUS) and branch vessel orifices (defined as interruptions or protrusions in the lumen contour which may have a variable shape).

### Data augmentation

To address the limited dataset size, extensive data augmentation was applied during training. The augmentation pipeline incorporated geometric and photometric transformations while preserving the physical realism of ultrasound imaging; (1) Random rotation (up to ±180 degrees) to simulate arbitrary catheter orientations during pullback, since IVUS catheters can rotate freely within the vessel lumen, (2) Intensity scaling (0.7– 1.3×) to mimic variations in ultrasound gain settings and tissue echogenicity across different anatomies, and (3) Speckle noise injection which adds multiplicative Rayleigh-distributed noise to simulate manufacturing variability in transducer imaging. Four augmentations were applied per labeled image, yielding an effective training set of approximately 18,000 augmented image-mask pairs.

### DeepLumen architecture design

DeepLumen is an encoder–decoder network that segments the aortic lumen and branch ostia from IVUS frames. In an ablation study (Fig. 2b), we compared four variants that extend a standard U-Net baseline with different strategies for enlarging the receptive field and incorporating spatial attention into the network:^63,34,49^

1. **Standard U-Net**: U-Net^64^ employs a symmetric encoder-decoder structure with skip connections that directly link encoder and decoder layers. Our implementation contains four encoder levels, batch normalization, and ReLU activations.
2. **ViT-UNet**: a small ViT encoder pretrained on ImageNet^65^ replaces the convolutional encoder while retaining the U-Net decoder structure. This hybrid CNN-Transformer architecture was included to evaluate whether transformer-based global attention could benefit IVUS segmentation.
3. **DRN-UNet:** To expand the receptive field of U-Net while preserving inductive biases inherent to CNNs, dilated residual nets^32^ (DRNs) were incorporated in the encoder pathway. Dilated convolutions insert gaps between kernel elements, exponentially increasing the receptive field without additional parameters that may yield overfitting behavior. Progressively increasing dilation rates (1, 2, 4, 8) were applied across encoder levels with residual shortcuts.
4. **CBAM-DRN-UNet (“DeepLumen”):** incorporates Convolutional Block Attention Modules (CBAM)^66^ on the skip connections between encoder and decoder, which sequentially applies channel attention (which features are important) and spatial attention (where to look), enabling the network to adaptively emphasize informative regions such as branch orifices while preserving inductive biases.

### DeepLumen cross-validation strategy

To prevent data leakage and maximize utilization of limited clinical datasets, a Leave-One-Patient-Out Cross-Validation (LOPOCV) protocol was employed. In IVUS imaging, sequential frames from the same patient are highly correlated both spatially and temporally. Standard random splitting would allow highly similar frames from the same patient to appear in both training and validation sets, artificially inflating performance metrics. LOPOCV addresses this by partitioning data at the patient level: for each of seven folds, the model is trained on six patients and validated on the omitted seventh patient. Reported Dice coefficient (DSC) metrics represent the mean and standard deviation across 7 folds from LOPOCV.

### DeepLumen training parameters

All architectures were implemented in TensorFlow 2.11 with the Keras API. TensorFlow and its XLA (Accelerated Linear Algebra) just-in-time compiler were selected, enabling local real-time inference at 15–30 Hz on a workstation laptop GPU (NVIDIA RTX A5500). Training required approximately 80 minutes per fold and inference times were ∼6ms per frame. Training was performed using the AdamW optimizer^67^ with the following hyperparameters: initial learning rate: α = 1 × 10^−4^, weight decay: λ = 0.01, Adam momentum parameters: β1 = 0.9, β2 = 0.99, batch size: 8, training duration: 20 epochs, He normal weight initialization, momentum: 0.9, dropout: rate 0.2, L2 weight regularization: λ = 10^−4^.

The loss function combines three complementary objectives to handle class imbalance and improve boundary localization:

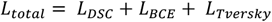

where the Dice Similarity Coefficient (DSC) loss emphasizes overlap between the network prediction and ground truth:

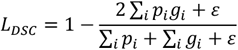

the Binary Cross-Entropy (BCE) loss is a pixel-wise classification loss that accounts for prediction confidence:

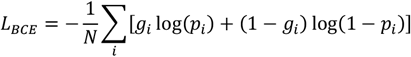

and the Tversky loss^68^ addresses class imbalance by penalizing false positives and false negatives asymmetrically:

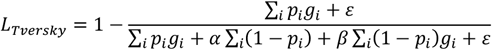

where *p*_*i*_ and *g*_*i*_ denote predicted and ground truth probabilities for pixel *i*, ε = 10^−7^ is a smoothing term, and α = 0.3, β = 0.7 are used to weight false positives and false negatives respectively. As described previously, a robust correspondence algorithm for the non-rigid registration pipeline was developed to filter false positives outputted by the segmentation network. Therefore, the asymmetric weighting in Tversky loss (β > α) prioritizes recall over precision, to ensure false negatives for the branch detections do not occur.

### DeepLumen in vivo fine-tuning details

To reduce the cost of in vivo animal studies while increasing dataset size, we collected IVUS pullback imaging from 5 female ex vivo porcine aortas and 3 male in vivo ovine subjects. The combined large animal dataset comprised approximately 1,700 images (∼250 labeled images per animal pullback). We performed fine-tuning on animal datasets to adapt to subtle differences between in vitro and in vivo imaging, which can arise from factors such as differences in the acoustic of hydrogel and native aortic tissue and the presence of the vena cava and abdominal organs not replicated in benchtop setups. During fine-tuning, the decoder layers were frozen while only the encoder remained trainable, to allow adaptation to domain-specific low-level features, while preserving the learned mapping for latent representations at the U-Net bottleneck.

### 3D intraoperative IVUS-EM reconstruction

To conduct 3D mapping of the aorta, segmented points from IVUS image must be transformed to the world coordinate system according to:

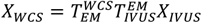

where *X*_*IVUS*_ are the segmented points in the 3D world coordinate system, 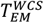is the measured six degree of freedom (6-DOF) EM pose, 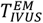is the calibration matrix defining the spatial offset between the EM sensor and IVUS imaging plane (determined using a calibration procedure described in Supplementary Text S1), and *XX*_*IVUS*_ are the segmented points in the ultrasound coordinate system. Signed Distance Functions (SDFs) are chosen to represent surfaces implicitly by storing the signed distance to the nearest surface boundary in a volumetric grid^35^. The reconstruction process segments each image into regions inside and outside the surface, computes per-pixel distances to boundaries, and integrates these per-frame SDFs into a global volumetric representation through weighted averaging^35^:

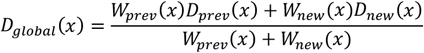

where *D*_*prev*_ (*x*) is the previous SDF value, *D*_*new*_(*x*) is the newly observed distance, and *W*(*x*) are confidence weights based on the number of observations in our case. This weighted averaging acts as a low-pass filter, suppressing noise while preserving geometric features across multiple observations. Surface meshes are extracted by identifying zero-crossing locations of the global SDF via the Marching Cubes algorithm^69^.

We represent the segmented vessel boundaries as a Euclidean Signed Distance Field (ESDF)^36^, computed by wave propagation from the segmented boundaries (Fig. S5). Unlike distance fields that assume line-of-sight (e.g. TSDF^35^), the ESDF accommodates the self-occlusion caused by curved vessel walls in IVUS. The resulting ESDF reconstructions form the intraoperative IVUS-EM aortic model used for subsequent fusion with preoperative CT.

### Semantic correspondence estimation with hidden markov model

We formulate correspondence as inference over vessel centerlines using a Hidden Markov Model (HMM) rather than dense surface matching (Fig. 3a), which reduces the search space and enforces sequential consistency along the vessel. This addresses three failure modes of standard nearest-neighbor and RANSAC matching under EM field distortion and anatomical deformation: convergence to local minima when geometric nearest neighbors diverge from true anatomical correspondences; false positives, when adjacent structures (e.g. inferior vena cava, hepatic vessels) are mistaken for aortic branches; and false negatives, when an oblique probe angle relative to the centerline causes the imaging plane to miss branch ostia (Supplementary Fig. S6). In our HMM formulation inspired by Chebrolu et al^40^, observations are the detected branch locations in IVUS and CT centerlines and hidden states are correspondence pairs *c*_*ij*_ linking branch *i* in IVUS to branch *j* in CT. ^40^We extract vessel centerlines from both IVUS and CT surfaces using Voronoi diagram-based skeletonization from the Vascular Modelling ToolKit (VMTK)^71^. Branch orifice candidates are identified from the raw segmented branch points using DBSCAN^72^, where the centroid of each cluster represents a candidate branch ostium.

The emission cost *Z(c*_*ij*_)quantifies the likelihood that IVUS branch *i* corresponds to CT branch *j* based on the node degree (the number of connected branches to a point) and geometric features (local aortic diameter and branch angle relative to centerline and other branches):

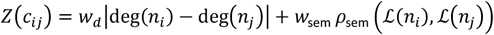

where deg(*n*) is the node degree, *L*(*nn*) encodes branch features (diameter and angle), *ρ*_sem_ is the cosine similarity computed based on these metrics, and *w*_*d*_, *w*_*sem*_ are weighting parameters. The weight terms were scaled such that both optimization terms were in a similar range, yielding *w*_*d*_ =100 and *w*_*sem*_ = 10.

The transition cost *Γ*(*c*_*ij*_, *c*_*kh*_) encodes the likelihood that correspondences *c*_*ij*_ and *c*_*kh*_ are mutually consistent, based on geodesic distance preservation between correspondence pairs and the number of matching branches:

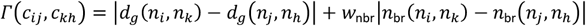

where *d*_*g*_ (*n*_*i*_, *n*_*k*_ ) is the geodesic distance along the centerline between nodes *i* and *k*, and *n*_br_(*n*_*i*_, *n*_*k*_ ) counts the number of branches between a correspondence pair set. The weight terms were scaled such that both optimization terms were in a similar range, yielding *w*_nbr_ = 10.

The optimal correspondence sequence is found via the Viterbi algorithm^73^, which efficiently computes:

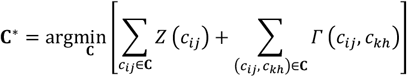

yielding the correspondence set **C**^∗^ that minimizes the total emission and transition costs. For reference, in cases with a large number of false positives (>7 spurious detections when matching 4 true abdominal branches), RANSAC’s probability of sampling a correct correspondence set within the permitted 50 iterations is 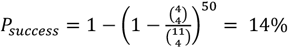

### Non-rigid deformation

Once robust correspondences are established between IVUS and CT via the HMM model, we deform the preoperative CT surface to match the intraoperative IVUS geometry. Rather than performing non-rigid registration of point clouds that are prone to self-intersections, we employ mesh-based registration that allows stiffness between mesh vertices to be tuned. Our approach builds on the non-rigid Iterative Closest Point (ICP) algorithm^41^, which minimizes an energy functional balancing alignment to the target geometry with smoothness of the deformation:

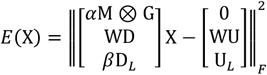

where **X** ∈ ℝ^4*V*×3^ are the stacked per-vertex affine transformations (rotation and translation), **M** ∈ ℝ^*E*×*V*^ is the edge incidence matrix encoding mesh topology, **G** = diag(1,1,1, γ) regularizes the affine transformations, **D** ∈ ℝ^*V*×4*V*^ applies transformations to homogeneous vertex coordinates, **W** ∈ ℝ^*V*×*V*^ are data term weights, **U** ∈ ℝ^*V*×3^ are nearest-neighbor correspondences on the target, **D**_*L*_ ∈ ℝ^*L*×4*L*^ applies transformations to landmark vertices, **U**_*L*_ ∈ ℝ^*L*×3^ are landmark target positions (derived from the HMM model), *α* controls stiffness, and *β* controls landmark influence. The stiffness term *α***M** ⊗ **G** penalizes differences in neighboring vertex transformations to enforce smooth, locally-affine deformations, as well as a landmark term *β***D**_*L*_ that anchors the registration to semantically-matched branch ostia identified by HMM correspondence estimation. The intuition for these terms is illustrated graphically in Supplementary Figure S7. To handle outliers in the data term, we employ a robust Huber kernel^74^ for computing correspondence weights:

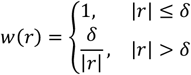

where *r* is the residual distance to the nearest target point and δ = 6 mm is the Huber threshold (the typical diameter of a branch).

We first apply non-rigid ICP to the vessel centerlines rather than full surface meshes. The centerline topology is encoded in the edge incidence matrix **M**, where each row corresponds to one centerline edge:

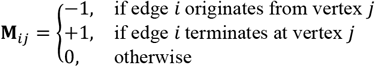

HMM-derived branch correspondences **C**^∗^ are embedded as landmark constraints in **D**_*L*_ and **U**_*L*_. The optimization proceeds through a gradual stiffness relaxation where *α* decreases from 200 to 1 over 20 iterations. At each iteration, the system is solved via sparse Cholesky decomposition^75^. Once the centerline is registered, we propagate its deformation to the full CT surface mesh using a mesh-following approach, where each surface vertex **v**_*i*_ is deformed according to a weighted combination of nearby centerline node transformations:

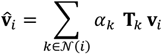

where *N*(*ii*) are the *K* nearest centerline nodes to vertex *i*, **T**_*k*_ ∈ *SE*(3) is the affine transformation computed for centerline node *k*, and *α*_*k*_ are normalized inverse-distance weights, similar to Sorkine et al^76^:

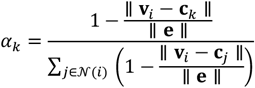

where **c**_*k*_ is the position of centerline node *k*, and ∥ **e** ∥ is the maximum edge length in the centerline to normalize distances. Finally, we apply standard non-rigid ICP directly to the deformed CT surface mesh to refine alignment with the IVUS-reconstructed surface.

### Pullback robot ECG gating details

The pullback increment is matched to the target mesh resolution (approximately 1.5 mm), ensuring that every axial cross-section of the aorta is imaged for a complete cardiac cycle, providing spatially-dense temporal information without requiring complex manual coordination of the catheter by the operator. For a typical adult aorta centerline length of approximately 38-42cm^77^ and a heart rate of 80bpm, the automated pullback image acquisition requires approximately 4 minutes. This acquisition time is similar to the time taken to perform a contrast injection with a power injector necessary for fluoroscopic guidance (3-5 minutes of preparation and administration^78^). 9 bins are used to gate the IVUS and EM transforms for creating separate 3D reconstructions.

### 4D motion capture validation

To quantitatively validate our 4D motion tracking accuracy, we compared IVUS-derived motion trajectories for three hydrogel phantoms against ground truth data acquired using a clinical 4D-CT scanner (Siemens SOMATOM, Siemens Healthineers). Patient-specific hydrogel phantoms were submerged in a water tank and connected to a pulsatile flow circuit driven by a HeartRoid pump (Fuyo Corportation) operating at 80bpm with 70mL stroke volume, replicating physiological cardiac output. An ECG simulator (Fluke Biomedical PS410) provided gating signals to the 4D-CT scanner, ensuring temporal synchronization. During 4D-CT acquisition, iodinated contrast (Omnipaque 350 mgI/mL, GE Healthcare) was injected into the phantom aorta via power injector (30mL at 2mL/s) with a 10-second delay. Imaging was triggered when contrast filled the aortic arch and continued for 15 seconds. The resulting phase-gated CT volumes were segmented using 3D Slicer^60^ to extract ground truth aortic surfaces at each cardiac phase. Given the labor-intensive nature of manual 4D-CT segmentation (approximately ∼10 hours for an expert per phantom to segment all phases), validation was performed on 3 hydrogel phantoms spanning a range of anatomical complexity (Patients 2, 4, and 6 from our dataset). Registration accuracy was quantified using bidirectional surface-to-surface distance. For each time point *t*, we computed the mean distance between corresponding surfaces:

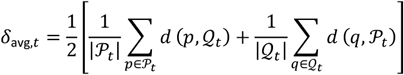

where *P*_*t*_ is the IVUS-registered CT surface and *Q*_*t*_ is the 4D-CT ground truth surface at phase *t*, and *d*(*p, Q*) denotes the minimum distance from point *p* to surface *Q*.

### Reconstruction of real-time catheter shape from EM sensors

Given the two EM poses and the known arc length *L* of the bending segment, we reconstruct the catheter shape assuming a constant curvature model. The bending angle *θ* is determined by employing an optimization algorithm that minimizes the distance between the predicted arc endpoint and the measured base position. The arc is parameterized in the tip coordinate frame, departing along the negative *x*-axis:

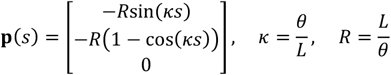

where *s* ∈ [0, *L*] is arc length and κ is curvature. The catheter points are then transformed to world coordinates via **p**_world_(*s*) = **R**_tip_ **⋅ p**(*s*) + **p**_tip_. For the proximal rigid segment between the femoral entry point **p**_0_ and the base of the steerable section **p**_1_, we employ quintic minimum-jerk spline interpolation to ensure smooth, physiologically realistic catheter trajectories:

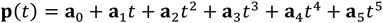

where *t* ∈ [0,1] and coefficients **a**_*i*_ are determined by boundary conditions: positions (**p**_0_, **p**_1_) and tangent directions (**v**_0_, **v**_1_) at the endpoints, with zero acceleration at both ends to minimize jerk.

### Real-time ECG-synchronized mesh update in augmented reality interface

The 4D registration framework synchronizes mesh deformation to the patient’s cardiac cycle in real time (Supplementary Figure S9). At each rendering frame visualized by Open3D, the current cardiac phase *αα* is computed from the ECG signal:

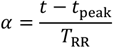

where *t* is the current time, *t*_peak_ is the timestamp of the most recent R-wave peak, and *T*_RR_ is the duration of the most recently measured R-R interval. Both *t*_peak_ and *T*_RR_ are updated continuously via real-time R-peak detection on the incoming ECG signal (or simulated ECG from the HeartRoid pump controller in the *in vitro* setting). The phase parameter *α* ∈ [0,1] maps to one of the *N* previously discretized cardiac phases (*N* = 9 as described earlier) from the 4DUS registration sequence. To obtain smooth motion between discrete phases, we identify the two adjacent phases *ϕ*_*i*_ and *ϕ*_*i*+1_ bracketing *αα* and apply linear interpolation of mesh vertex positions:

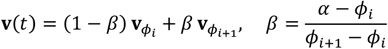

where **v**_*ϕi*_ and **v**_*ϕi*+1_ are vertex positions from the registered CT meshes at phases *i* and *i* + 1. This approach ensures that the rendered anatomy tracks the phantom’s instantaneous cardiac phase, automatically adapting to heart rate variability. Given the non-inferiority between static and dynamic motion compensation (as mentioned in Results section), a 3D printable rubber polymer TEPU 30A (Inkbit, Cambridge, USA) was used for the phantom material during static in vitro validation studies (Fig. 6), given its ease of manufacture. TEPU 30A provides similar ultrasound compatibility to hydrogels, and differs primarily in its mechanical compliance. In this regime, the rendered mesh remained fixed and the mesh update framework was not applied.

### Operator training

Prior to data collection, surgeons received a brief 5-minute orientation of the AR interface consisting of: (1) demonstration of the dual-view AR interface (global 3D and endoscopic views), (2) explanation of branch ostia visualization as colored ring targets, and (3) instruction on catheter articulation and guidewire advancement mechanics.

### IVUS fiducial integration with physician modified endograft planning

A patient-specific aneurysm phantom was created with a maximum diameter of 5.5cm (the threshold at which intervention is typically recommended^79^). The aneurysm is 10 cm in length, with a 24 mm aortic neck diameter at the planned proximal and distal landing zones. For our demonstration, we use a Cook Zenith Alpha^−^ Thoracic Endovascular Graft (ZTA2-P-28-155-W, Cook Medical, Bloomington, IN), which yields 17% oversizing for an adequate seal within our 2cm aortic neck diameter (within clinical guidelines of 10-20%^79^). The 155 mm length allows for proximal and distal landing zones of 2.5 cm each. We demonstrate how IVUS fiducial integration can be incorporated into Physician Modified EndoGraft (PMEG) workflow, which is commonly used to create a fenestrated graft given a non-fenestrated off the shelf EVAR template (Figure 7b). In this regime, surgeons analyze CT imaging to determine the optimal location of fenestrations on the graft. The graft template was unsheathed from its delivery system and longitudinal and circumferential measurements of the branch orifices derived from the preoperative CT scan were projected onto the graft fabric using a marker. Fenestrations approximately 6 mm in diameter were created using electrocautery to accommodate standard 6-8 Fr stent delivery systems^80,81^. Tungsten beads (0.8 mm diameter) were affixed to the inner graft surface immediately proximal to each fenestration using cyanoacrylate adhesive, though could be sutured clinically. This proximal offset ensures that beads do not obstruct the fenestration opening while still being observable with IVUS. Given the known fenestration radius (3 mm) and consistent proximal bead placement, the fenestration center can be geometrically inferred from the bead position detected via IVUS. Finally, the modified graft was re-crimped onto the deployment catheter using a hand-crank crimper (Model RVS, Blockwise Engineering, San Jose, CA) and gradually re-sheathed within the Zenith Alpha delivery system with assistance from retention sutures. To deploy the graft, the catheter delivery system was introduced via femoral access and advanced to the aneurysm under fluoroscopic guidance. The graft was partially unsheathed and positioned both longitudinally and rotationally to align the fenestrations to their corresponding branch orifices. The tungsten beads serve as radiopaque markers to aid orientation of the fenestrations until the graft is fully deployed (Supplementary Figure S12).

### FEVAR augmented reality computational pipeline details

Prior to FEVAR deployment, the aortic aneurysm geometry is mapped and an IVUS-CT registration is obtained. After obtaining the registration and following graft deployment, an IVUS-EM pullback through the graft lumen is performed to create an approximate 3D ESDF reconstruction of the inner surface and to localize the tungsten beads. A prior parametric virtual model of the graft was constructed from mesh primitives including tubular geometry to represent the graft fabric and 3D sinusoidal wire frames to represent the stent struts. The exact dimensional specifications for the virtual parametric graft model were obtained from the Cook Medical Zenith Alpha reference guide (Cook Medical, Bloomington, IN, USA). This parametric non-fenestrated virtual EVAR model was then registered to the 3D ESDF graft reconstruction. For our proof-of-concept demonstration, bead positions were identified via manual annotation in the IVUS image by clicking bright spots in the GUI. This approach is feasible given we only need to click four discrete points per FEVAR graft. We note this manual step could be automated using a modified DeepLumen architecture trained to segment an additional “bead” class alongside lumen and branch boundaries. Fenestrations were created in the virtual model by extruding 6 mm diameter cylinders normal to the graft surface and performing a boolean subtraction.

### Ovine preoperative CT imaging

Preoperative contrast-enhanced CT angiography was performed 48 hours prior to the interventional procedure to allow for image segmentation and surgical planning (Fig. 8a). All procedures were approved under IACUC protocol and conducted in accordance with institutional animal care guidelines. We selected an ovine (sheep) model given ovine aortic dimensions are more closely matched to human anatomy than porcine models. Animals were induced with propofol (6-8 mg/kg IV bolus) and maintained under general anesthesia with isoflurane (1.5-2.5% inhalational). Internal jugular venous access was established under ultrasound guidance (Siemens ACUSON, Siemens Healthineers) to enable powered contrast injection. A scout CT scan (GE Discovery CT750 HD, GE Healthcare) defined the imaging field from the aortic brachiocephalic trunk to the femoral arteries, encompassing the entire thoracoabdominal aorta and major branch vessels. ECG monitoring (Dash 4000, GE Healthcare) was established via four-lead placement (right arm, left arm, right leg, left leg), with baseline heart rates of 90 bpm (Sheep 1), 87 bpm (Sheep 2) and 103 bpm (Sheep 3). Animals were mechanically ventilated to enable a 15-second breath hold during image acquisition, minimizing respiratory motion artifacts. Contrast-enhanced imaging was triggered when Hounsfield unit (HU) density in the aortic arch reached 125 HU following injection of iodinated contrast (Omnipaque 350 mgI/mL, GE Healthcare; 2mL/kg at 5mL/s). Helical acquisition proceeded for 15 seconds with 0.625 mm slice thickness and ECG gating for cardiac phase synchronization. Animals were recovered following CT acquisition and housed for 48 hours prior to the interventional procedure. Following scan acquisition, aortic segmentation was performed using 3D Slicer^60^, requiring approximately 2 hours per animal.

**Fig. 8.**
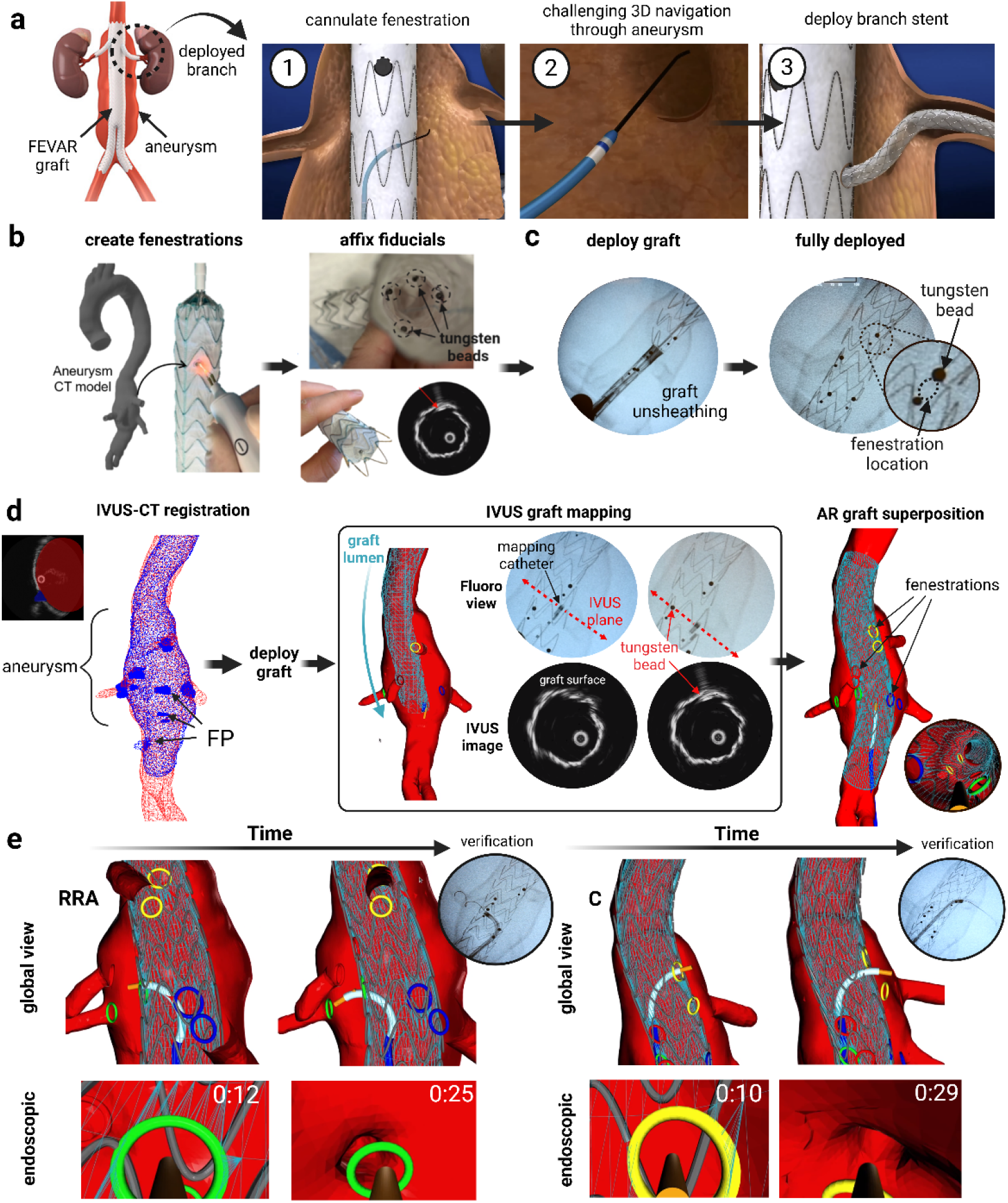
Platform demonstration in vitro for fenestrated endovascular aneurysm repair (FEVAR). a. A fenestrated endograft is deployed when an abdominal aortic aneurysm (AAA) involves the renal arteries, requiring fenestrations (i.e., openings) in the graft fabric to maintain perfusion to visceral organs. A steerable catheter is navigated from within the endograft lumen through a fenestration and into the aneurysm sac to deploy a bridging stent (panel adapted from “FEVAR robotics,” Hansen Medical, Mountain View, CA). **b**. Physician-modified endograft (PMEG) preparation workflow adapted for AR guidance. The graft is unsheathed and fenestration positions are marked based on CT scan information. Tungsten beads are affixed to the graft to serve as IVUS-visible fiducials for intraoperative fenestration localization. **c**. The fenestrated deployment system is positioned within the aneurysm under fluoroscopic guidance, with fenestration boundaries (indicated by dashed lines) inferred from the spacing between adjacent tungsten beads. **d**. AR-guided FEVAR computational pipeline. The AAA is mapped using IVUS–EM and registered to preoperative CT anatomy via the non-rigid fusion framework described previously. After graft deployment, a second IVUS pullback is performed through the graft lumen, where tungsten beads appear as hyperechoic spots and serve as fiducials for precise 3D localisation of the graft shape and fenestration positions. The reconstructed FEVAR graft geometry is then superimposed onto the registered AAA anatomy within the AR display. **e**. Qualitative comparison of target vessel cannulation under fluoroscopic and AR guidance for the right renal artery (RRA) and celiac (C) fenestrations. When fenestration-to-branch misalignment is present, the catheter must be redirected within the aneurysm sac to locate the target ostium. Supplementary Video S7 provides a visual demonstration of the computational workflow and augmented reality interface.

### Ovine surgical preparation

Three male Dorset sheep (48kg, 52kg, and 54kg) were selected to ensure femoral artery diameters sufficient for 12 Fr sheath access required by the hybrid IVUS-EM mapping catheter. On the day of the procedure, animals were re-anesthetized using the same protocol detailed above and maintained on mechanical ventilation. Percutaneous femoral arterial access was established using the Seldinger technique^82^ under ultrasound guidance (Supplementary Figure S13). Based on preoperative CT, the optimal access site was identified at the level of the inguinal ligament. Transabdominal ultrasound (7.5 MHz linear probe, Siemens ACUSON, Siemens Healthineers) visualized the femoral artery and vein, with the artery distinguished by, (1) arterial pulsatility, (2) resistance to compression (veins collapse under probe pressure), and (3) medial position relative to the femoral vein. A 21-gauge micropuncture needle was advanced into the femoral artery at a 45° angle under real-time ultrasound visualization. After the 0.018” guidewire was introduced, the needle was exchanged for a dilator to progressively dilate to 12 Fr. A 12 Fr hemostatic sheath (Gore Dryseal, Cook Medical) was placed, providing sealed access for catheter exchanges throughout the procedure. Finally, the hybrid mapping catheter was advanced over a 0.014” hydrophilic guidewire (MAILMAN, Boston Scientific) from the femoral sheath into the thoracic aorta under fluoroscopic and IVUS guidance. The catheter tip was positioned superior to the celiac artery, the catheter was loaded into the automated pullback robot, and an ECG-synchronized pullback was initiated from the thoracic aorta down to the aortic bifurcation. Following data collection, the IVUS frames, EM poses, and ECG timestamps were ECG-gated and registered to the segmented CT anatomy using the previously described non-rigid fusion framework. After obtaining the CT-IVUS registration, TVC was performed on all branches under two conditions: (1) AR-guided platform, and (2) fluoroscopy guidance with digital subtraction angiography.

### Ovine study digital subtraction angiography

To establish a performance baseline, target vessel cannulation was performed under standard fluoroscopic guidance with digital subtraction angiography (DSA) roadmapping (Supplementary Figure S14). In this regime, a 5 Fr pigtail catheter was positioned in the descending thoracic aorta, and iodinated contrast (Omnipaque 350 mgI/mL) was injected during fluoroscopic acquisition. The resulting angiogram can be overlaid as a transparent map on live fluoroscopy imaging to act as a roadmap. With this DSA roadmap, the surgeon attempted to cannulate all four abdominal branches (celiac, SMA, left renal, right renal) using the steerable catheter. Successful cannulation was defined as guidewire advancement into the branch without coiling within the aortic lumen as verified using fluoroscopy.

### Statistical analyses

Reported metrics represent mean ± standard deviation. Segmentation performance across architectures and correspondence estimation methods were compared using one-way ANOVA with Tukey post hoc tests for pairwise comparisons. TVC performance metrics (cannulation time, fluoroscopy exposure, and number of attempts) between AR and fluoroscopic guidance were compared using two-sided paired t-tests. NASA Task Load Index subscale scores were compared using paired t-tests. Statistical significance was defined as p < 0.05.

## Acknowledgements

This work was supported by the MIT-MGB health and life sciences collaborative grant. T.D. acknowledges the financial support through a scholarship from MIT School of Engineering MathWorks Fellowship. We thank Becky Woodcock, Kichang Lee, Moshe Yaghoubian, Andres Alfonso Lezama (Massachusetts General Hospital) for help with animal testing. We also thank Marina McDonald (Massachusetts General Hospital) for help with preoperative CT ovine imaging. We thank Marc Schermerhorn and Patric Liang (Beth Israel Deaconness Medical Center Boston) for insightful discussion and constructive feedback on the developed system.

## Contributions

T.D. conceived the research idea and high level platform design, designed research methodology and aims, designed the hybrid IVUS-EM catheter, spatial calibration procedures, interprocess communication framework, DeepLumen framework, static and dynamic non-rigid registration framework, 4DCT validation protocol, pullback robot high-level design and software, AR FEVAR PMEG and computational workflow, graphical user interface, performed manual segmentation of aortic CT for ground truth data, conceived the high-level fabrication workflow for the hydrogel in vitro phantoms, designed experimental preclinical validation procedures (*in vitro*, FEVAR, *in vivo*), led preclinical validation studies, performed data analysis, and wrote the manuscript. D.Q. refined and implemented the hydrogel phantom fabrication workflow for hydrogel *in vitro* phantoms, designed the water tank, and fabricated the pullback robot hardware. B.A. provided clinical feedback on platform user interface, led IACUC protocol approval, and participated in *in vitro* studies and large animal surgeries. B.S. participated in *in vitro* studies and large animal surgeries. E.K.R. contributed to development of the *in vitro* hydrogel phantom fabrication pipeline. B.K. and J.T. participated in *in vitro* studies. E.T.R. supervised the project and provided feedback on the manuscript.

## Competing Interests

T.D, D.Q, B.A, and E.R. are listed as inventors of a patent on the presented ultrasound-driven navigation platform. The other authors declare that they have no competing financial interests.

## Data Availability

All data supporting the findings of this study are available within the article and its supplementary files but can be found in Supplementary Data 1. We release our in vitro and in vivo 3D datasets (https://doi.org/10.5281/zenodo.20737791) to support the development of registration algorithms for IVUS-based navigation. Moreover, we release labeled IVUS imaging (https://doi.org/10.5281/zenodo.20752361) of the aortic lumen and branch for for researchers developing AI algorithms for IVUS interpretation. Any additional requests for information can be directed to, and will be fulfilled by, the corresponding authors.

## Notes

### Competing Interest Statement

The authors have declared no competing interest.

