## Supplementary Information for "Learned ultrasound segmentation and deformable CT fusion for augmented reality endovascular surgery"

BioRxiv Preprint

#### Contents

Supplementary Text S1–S10

Supplementary Figures S1–S14

Supplementary References

#### Additional supplementary files

Supplementary Movie 1–7

Supplementary Data 1

### Supplementary Text

#### S1 Spatial Calibration of IVUS-EM

A key requirement for 3D mapping with an IVUS-EM system is accurate determination of $T^{EM}_{IVUS}$, the spatial offset between the ultrasound imaging plane and the EM sensor frame. Since the EM sensor can be mounted at any circumferential position, rotational configuration, and translational location relative to the IVUS probe, this offset must be explicitly calibrated. This is analogous to the hand-eye calibration problem in robotics [1], where the spatial relationship between a camera and a robotic end-effector is determined from a set of known reference points (usually obtained from a checkerboard pattern) [2].

For our application, we employ a string phantom fabricated with a known geometric arrangement of thin wires submerged in water (**Figure S1**). An EM sensor is embedded within the phantom, allowing the ground truth pose of the string phantom to be determined in 3D space. Under ultrasound imaging, each wire cross-section appears as a localized bright spot whose centroid can be readily segmented and localized in the image plane. While least-squares approaches have been proposed for automated calibration parameter estimation [3], these methods are susceptible to local minima in the presence of segmentation outliers. Instead, we developed an interactive calibration GUI that enables real-time visualization and adjustment of the transformation parameters in 3D space, allowing the operator to directly inspect alignment between segmented wire centroids and their known ground truth positions. This real-time feedback significantly simplifies the calibration procedure.

#### S2 Hand-crafted Segmentation Comparison

Hand-crafted segmentation algorithms that rely on predictable edge detection with per-patient parameter tuning can be unreliable across the full spectrum of clinical anatomies and imaging conditions. The necessity of a deep learning approach was validated by benchmarking DeepLumen against a hand-crafted lumen segmentation algorithm employing raycasting, followed by RANSAC-based ellipse fitting and

B-spline extrapolation. This is consistent with state-of-the-art hand-crafted IVUS segmentation algorithms[4, 5]. DeepLumen significantly outperformed this classical baseline as shown in **Figure S2**, justifying the adoption of a learned segmentation model.

#### S3 Benchmarking against Coherent Point Drift (CPD)

Widely employed non-rigid registration pipelines such as Coherent Point Drift (CPD) [6] often utilize soft correspondence schemes which do not exploit the semantic structure of the geometry and therefore remain vulnerable to local minima in the presence of numerous outliers. We benchmarked our semantic non-rigid registration framework against CPD. Our pipeline reduced branch localization error from $10.51 \pm 6.45$ mm to $1.76 \pm 0.98$ mm (n=7 in vitro phantoms, p $= 1.19 \times 10^{-8}$).

#### S4 Hyperparameter sensitivity

Our non-rigid registration pipeline is largely insensitive to the choice of stiffness weight $\alpha$ and landmark weight $\beta$. As shown in **Figure S8** registration accuracy is preserved across roughly four orders of magnitude in each ($\alpha \in [0.01, 1000]$, $\beta \in [0.1, 1000]$).

#### S5 NASA Task Load Index Assessment Details

Cognitive demand was assessed using the NASA Task Load Index (NASA-TLX) [7] which is a widely used, multidimensional questionnaire used in human factors research that measures a person's perceived mental workload after performing a task or working with a system. Surgeons completed the questionnaire after the experiments, rating mental demand, physical demand, temporal demand, performance, effort, and frustration on 100-point scales (lower scores indicate reduced workload). AR guidance significantly reduced cognitive workload across four of six dimensions for the 4 surgeons, namely: Mental demand, representing the cognitive effort required for navigation and decision-making, decreased by 30% (AR: $56.3 \pm 11.1$, X-ray: $80.0 \pm 0.0$; $p = 0.023$, paired t-test; **Figure S10**); Temporal demand, or perceived time pressure, was reduced by 50% (AR: $33.75 \pm 17.5$, X-ray: $70.0 \pm 20.4$; $p = 0.043$); Effort required for task completion decreased by 40% (AR: $47.5 \pm 17.1$, X-ray: $78.8 \pm 14.4$; $p = 0.038$);

and frustration was reduced by 53% (AR: $32.5 \pm 18.9$, X-ray: $68.8 \pm 21.0$; $p = 0.038$).

In contrast, physical demand showed no significant difference between conditions (AR: $40.0 \pm 32.4$, X-ray: $58.8 \pm 23.9$; $p = 0.395$), which is expected given that both modalities use identical catheter hardware for target vessel cannulation. Perceived performance, which is an assessment of one's own perceived performance, did not differ significantly (AR: $25.0 \pm 19.1$, X-ray: $57.5 \pm 20.6$; $p = 0.069$). This may reflect surgeons' limited familiarity with the AR platform at the time of testing, despite having achieved objective performance improvements (faster times, fewer attempts).

#### S6 Learning Curves

To assess skill acquisition with the AR platform, we analyzed cannulation time trajectories across the seven sequential phantoms for each surgeon. While some variation can be attributed to inter-phantom variability (for example, Phantom 5 exhibited relatively straight anatomy that was easy to cannulate even under single-plane fluoroscopy), a clear downward trend in AR cannulation times is evident across the experimental sequence (**Figure 6c**). Averaged across all four residents, AR cannulation times decreased from 27 seconds (Phantom 1) to 12 seconds (Phantom 7), a 56% improvement (slope: $-2.8$ s/phantom). Fluoroscopy times showed minimal systematic improvement (slope: $-0.3$ s/phantom), consistent with surgeons' extensive prior experience with X-ray guidance (minimum 3 years for all participants). Notably, the standard deviation in AR times decreased from 8.6 seconds (Phantom 1) to 3.2 seconds (Phantom 7), indicating not only faster performance but also more consistent, streamlined execution. These results are particularly encouraging given all participants had 3+ years of fluoroscopy experience prior to the experiments, which suggests that the AR interface is highly learnable despite its novelty.

#### S7 Impact of 4D Motion Compensation on Navigation Performance

To quantify the benefit of dynamic motion tracking over static registration, we compared cannulation performance using 4D motion-compensated AR guidance versus conventional 3D static guidance in the 3 patient-specific phantoms validated with 4D-CT ground truth. Both conditions employed identical pulsatile flow (80 bpm, 70 mL stroke volume via HeartRoid pump) and task protocols (cannulate all four branches as rapidly as possible). The only experimental variable was whether the AR platform displayed a dynamically-updating 4D registration or a time-invariant 3D reconstruction. In the 3D static condition, the rendered mesh remained fixed at the end-diastolic configuration (phase 0) regardless of the actual phantom motion, representing the current clinical standard where preoperative CT anatomy is assumed stationary during the procedure.

**Figure S11** presents cannulation performance under 3D static and 4D motion-compensated guidance across the three validated phantoms ($N = 48$ total cannulations: 4 branches $\times$ 3 phantoms $\times$ 4 operator runs). Mean cannulation time was numerically lower with 4D motion compensation (4D: $21.8 \pm 13.2$ s, 3D: $26.0 \pm 11.8$ s), representing a 16% reduction, though this difference did not reach statistical significance ($p = 0.105$, paired t-test). Given the smaller motion scale in the abdominal aorta, surgeons can successfully cannulate branches using time-averaged static anatomy without requiring real-time motion compensation.

Nonetheless, the 4D framework presented may be more important in anatomical regions exhibiting larger cardiac-driven motion. As discussed previously, the thoracic aorta and aortic arch undergo displacements exceeding 10–14 mm during systole [8]. Procedures such as transcatheter aortic valve replacement (TAVR) and thoracic endovascular aortic repair (TEVAR) where cardiac-driven motion is substantial would likely derive significant benefit from motion-compensated visualization.

#### S8 Phantom Material Selection

To replicate native aortic characteristics *in vitro*, the phantom material must satisfy three key requirements: (1) mechanical compliance similar to native aortic tissue to enable realistic deformation under pulsatile flow, (2) low surface friction to simulate authentic catheter–vessel interactions, and (3) an acoustic impedance matching that of human tissue to reproduce clinically representative ultrasound image quality and artifacts. Hydrogel materials have gained widespread use in medical device research due to their biocompatibility and ability to closely mimic the mechanical and acoustic properties of soft biological tissues [9]. Polyvinyl alcohol (PVA) hydrogel, in particular, exhibits acoustic impedance closely matched to human aortic tissue. Native aortic tissue has a density of approximately 1060 kg $m^{-3}$ and sound speed of 1540 m $s^{-1}$, yielding an acoustic impedance of $1.63 \times 10^6$ rayl [9, 10, 11]. PVA hydrogel formulations achieve comparable values[12, 13] (density: 1020–1050 kg $m^{-3}$; sound speed: 1520–1560 m $s^{-1}$; impedance: 1.55–$1.64 \times 10^6$ rayl), enabling realistic ultrasound image formation. Additionally, PVA hydrogels can be tuned through freeze–thaw cycling to exhibit physiologically relevant compliance [14] (Young's modulus: 50–400 kPa), comparable to native aortic tissue at physiological pressures [15] (50–300 kPa), and possess inherently low surface friction due to their high water content and surface hydrophilicity. Mix *et al.* have employed PVA hydrogel to fabricate idealized aortic phantoms for ultrasound validation [16]. However, these approaches cannot capture the anatomical complexity, branch vessel configurations, and tortuosity characteristic of patient-specific aortic anatomy visible in clinical CT scans.

#### S9 Random Sample Consensus (RANSAC) for Non-Rigid Registration

Random Sample Consensus (RANSAC) estimates transformation models by iteratively sampling minimal correspondence sets of 3 valid point pairs and counting inliers for rigid registration. Extending RANSAC to dense non-rigid problems without modification where every mesh vertex can move independently is typically computationally intractable. Our centerline-based representation enables a practical RANSAC baseline by sampling correspondence hypotheses only for branch landmarks and interpolating intermediate correspondences along geodesic paths.

#### S10 User Interface Design

The augmented reality (AR) interface was built on Open3D and ROS. A workstation laptop running the augmented reality guidance interface is projected to a monitor positioned adjacent to the fluoroscopy display. Branch ostia are highlighted as colored rings positioned at their registered locations from the CT-IVUS fusion, serving as navigational targets. An operator (e.g., imaging technician or surgical assistant) can interactively manipulate the camera viewpoint using mouse or joystick controls, negating the need for continuous C-arm repositioning typical of fluoroscopy. The continuous 3D visualization enables surgeons to plan direct trajectories to target branches rather than relying on iterative trial-and-error navigation. As described by one participant: "I initially get my bearings by looking at the global 3D view, and then refine the final tip alignment by looking at the endoscopic view."

### Supplementary Figures

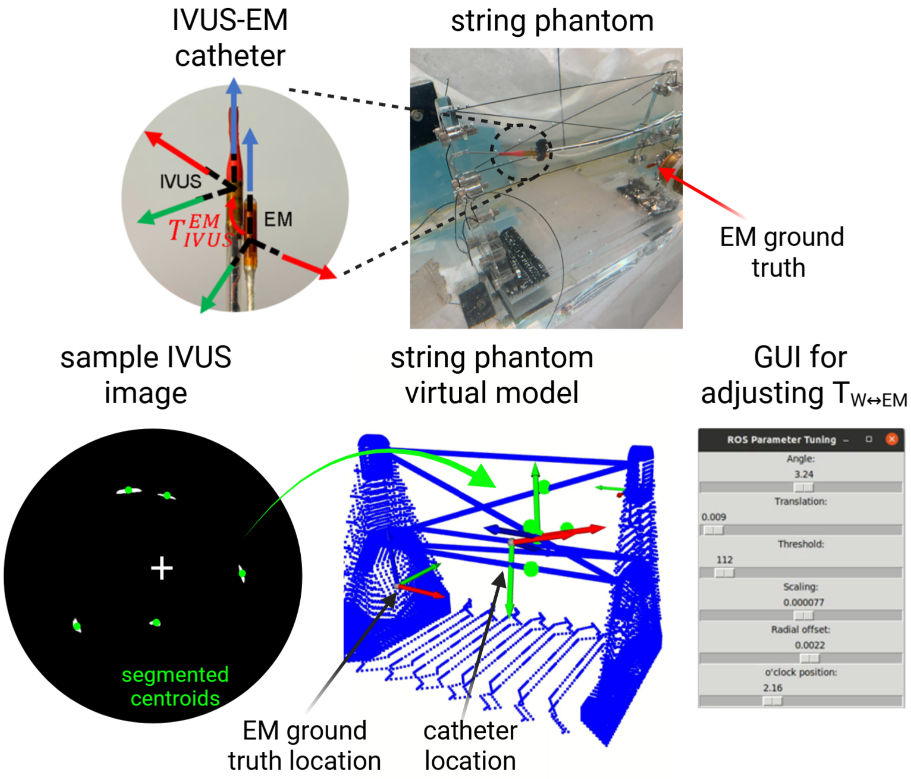


**Figure S1: Solving the Hand-Eye Calibration Problem in IVUS-EM Mapping.** A custom string phantom was fabricated for IVUS imaging to enable unique triangulation of the imaging plane pose from a single IVUS image. An EM sensor embedded within the phantom provides ground truth positions of the known wire geometry. When the IVUS catheter is positioned parallel to the angled wires, each wire cross-section appears as a distinct bright spot that can be segmented and transformed to 3D space using a candidate set of calibration transformation $T_{IVUS}^{EM}$. A real-time GUI enables interactive adjustment of the transformation parameters until strong alignment is achieved between the known phantom geometry and the transformed segmented points.

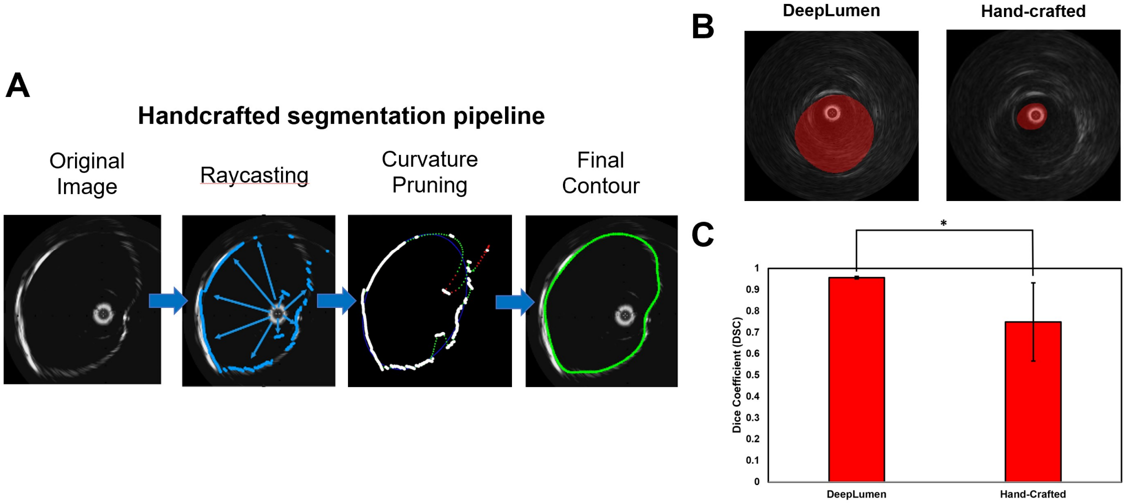


**Figure S2: Benchmarking of DeepLumen Against a Hand-Crafted Lumen Segmentation Algorithm.** (**A**) Hand-crafted segmentation pipeline: raycasting is performed radially outward from the probe to obtain a preliminary lumen contour. Outlier points (defined as those exceeding a predefined inlier distance from a fitted ellipse or exhibiting high curvature) are pruned via RANSAC. A B-spline is then fitted to the remaining points to produce the final contour. (**B**) Qualitative comparison demonstrating that DeepLumen is robust to reverberation artifacts, which manifest as a spurious secondary contour proximal to the probe and do not represent the true lumen boundary. (**C**) Quantitative lumen segmentation performance across both methods, demonstrating that DeepLumen significantly outperforms the hand-crafted approach. Error bars denote ± SD. Asterisks indicate statistical significance at $p < 0.05$ (Welch's $t$-test).

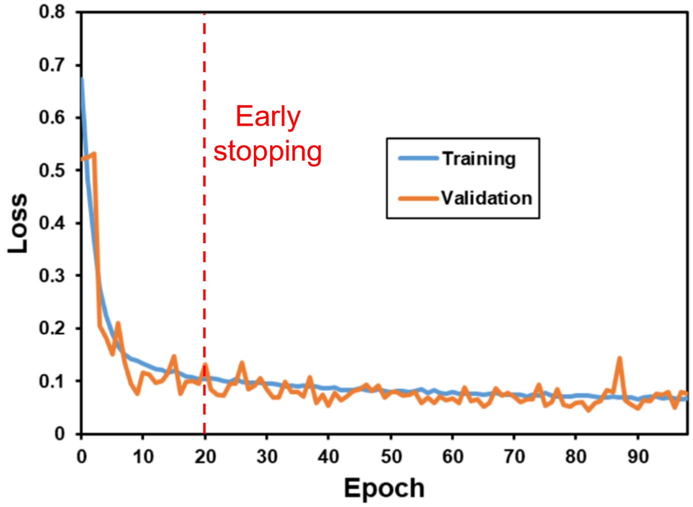


**Figure S3: Sample loss curve during DeepLumen training.** Training loss (blue) and validation loss (orange) for a representative LOPOCV fold. Both curves plateau after approximately 20 epochs, with validation loss showing no sustained improvement beyond this point. Continued training beyond this threshold would risk overfitting to the training set.

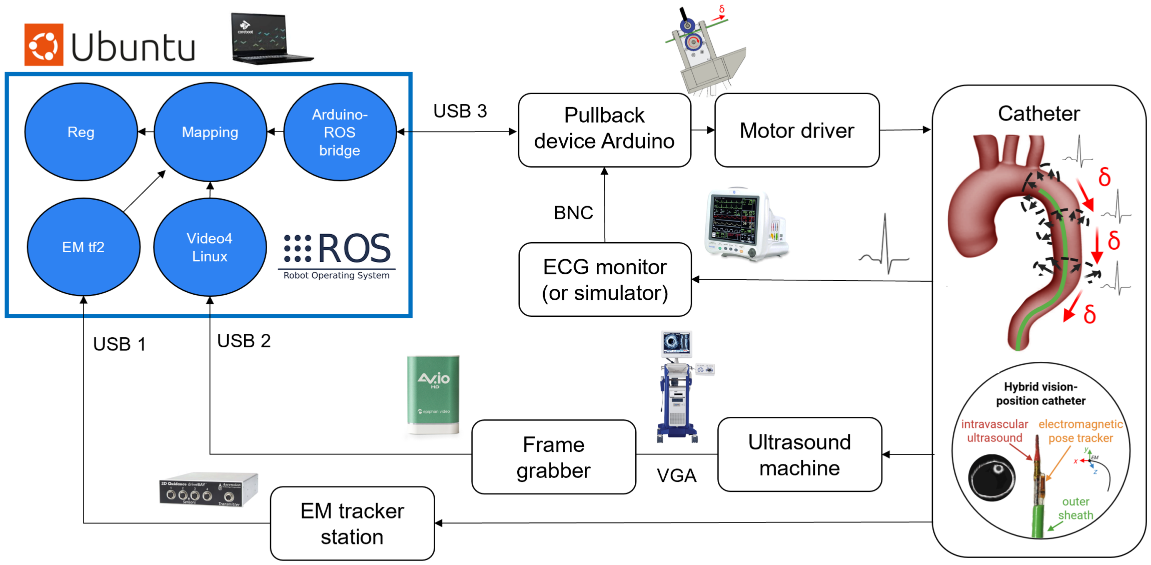


**Figure S4: Intraoperative IVUS Mapping Inter-Process Communication Framework.** A workstation running Ubuntu Linux executes the Robot Operating System (ROS) middleware for real-time inter-process communication. USB interfaces stream data from the EM tracker, IVUS console, and pullback robot to the host computer where ROS nodes are running (highlighted in blue). The ECG monitor (or ECG signal simulator) coordinates pullback motion and provides cardiac gating signals for temporal image synchronization. A frame grabber digitizes the IVUS video output for real-time image streaming to the ROS pipeline.

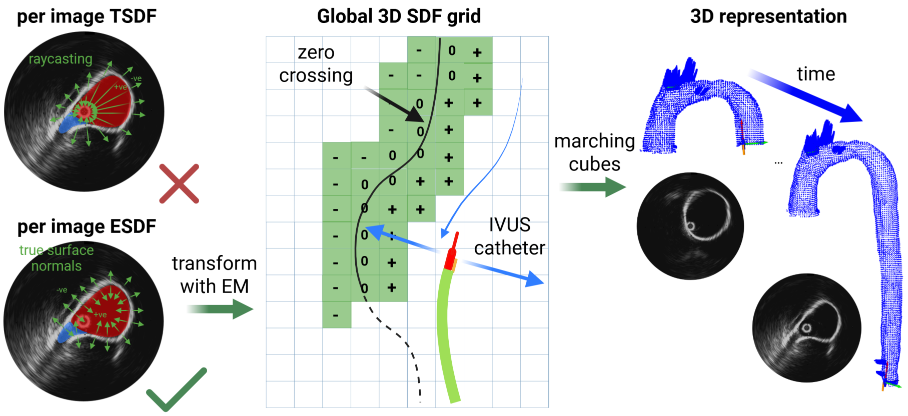


**Figure S5: Real-Time IVUS-EM 3D Reconstruction Pipeline.** The Euclidean signed distance field (ESDF) computes distances via wave propagation from segmented boundaries [17], providing a more accurate measured of SDF than raycasting-based TSDF methods [18]. Per-frame ESDFs are integrated into a global volumetric representation through weighted averaging, which filters measurement noise while preserving geometric features. The zero-level set of the global SDF represents the aortic surface and is extracted as a triangulated mesh via the Marching Cubes algorithm [19]. The resulting mesh vertices can be represented as a point cloud that serve as a target surface for CT-IVUS registration.

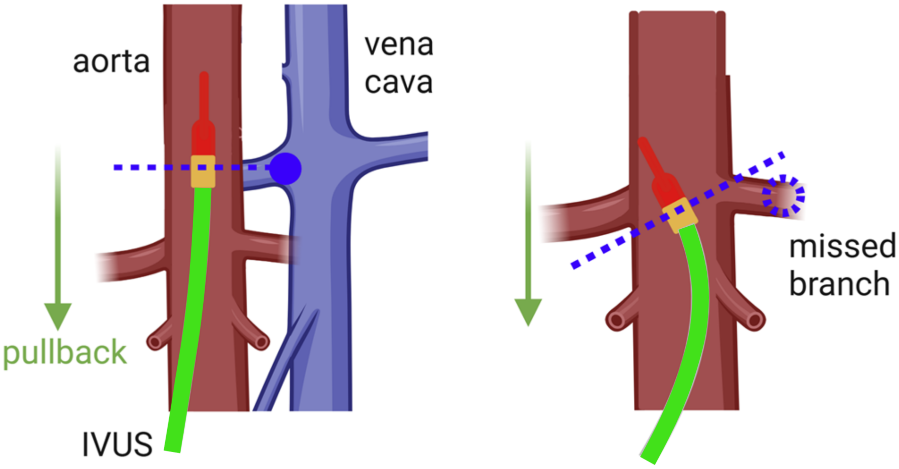


**Figure S6: False Positive and False Negative Branch Detections During IVUS Pullback.** False positives occur when adjacent anatomical structures (e.g., inferior vena cava, hepatic vessels) are confused for aortic branches. False negatives arise when the IVUS probe is angled obliquely to the centerline, causing the imaging plane to miss branch ostia.

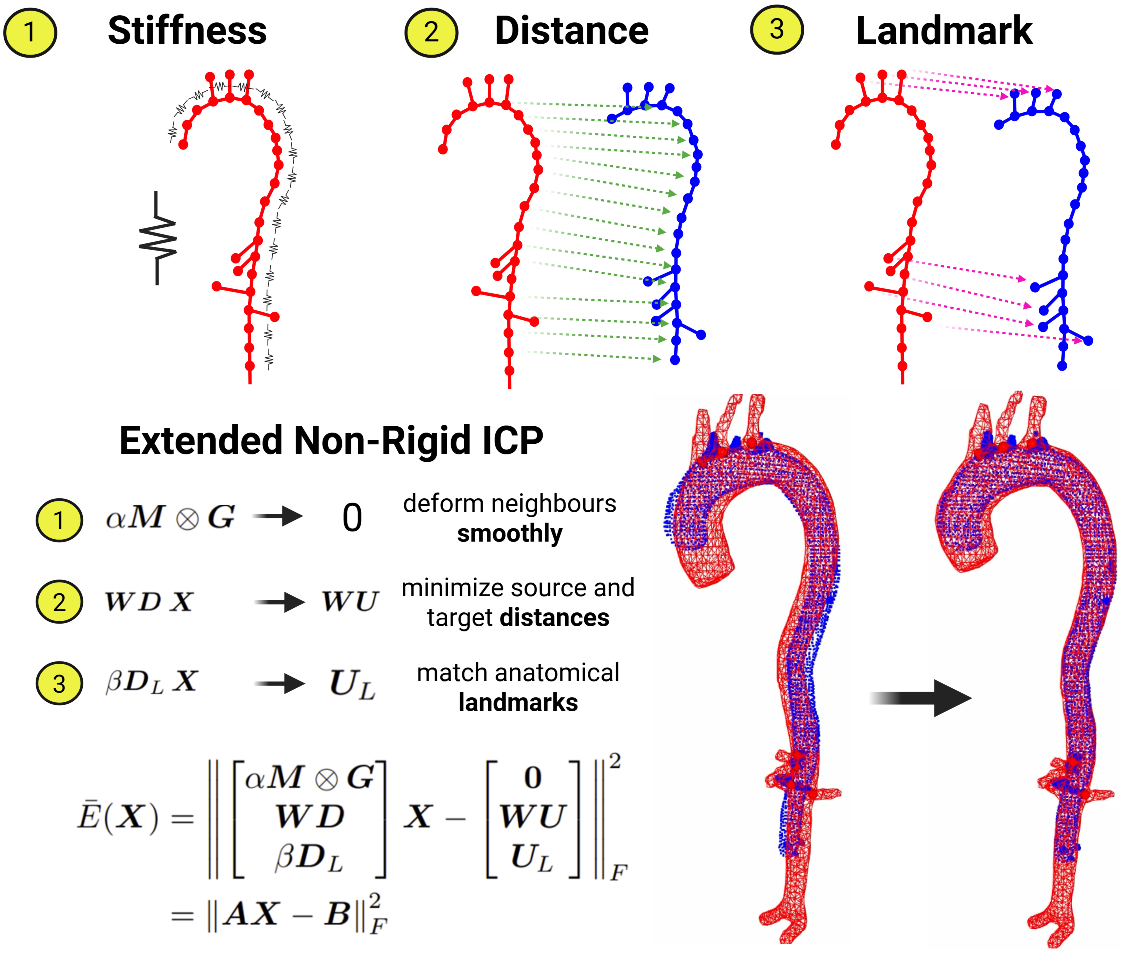


**Figure S7: Non-Rigid Deformation of CT scan onto IVUS data.** The optimization contains 3 primary components - (1) a stiffness term that minimizes relative displacements between neigbouring nodes during the deformation, (2) a distance term that attracts the source surface to nearest neighbours on the target surface, and (3) a landmark term that attempts to match known corresponding points derived from the HMM model.

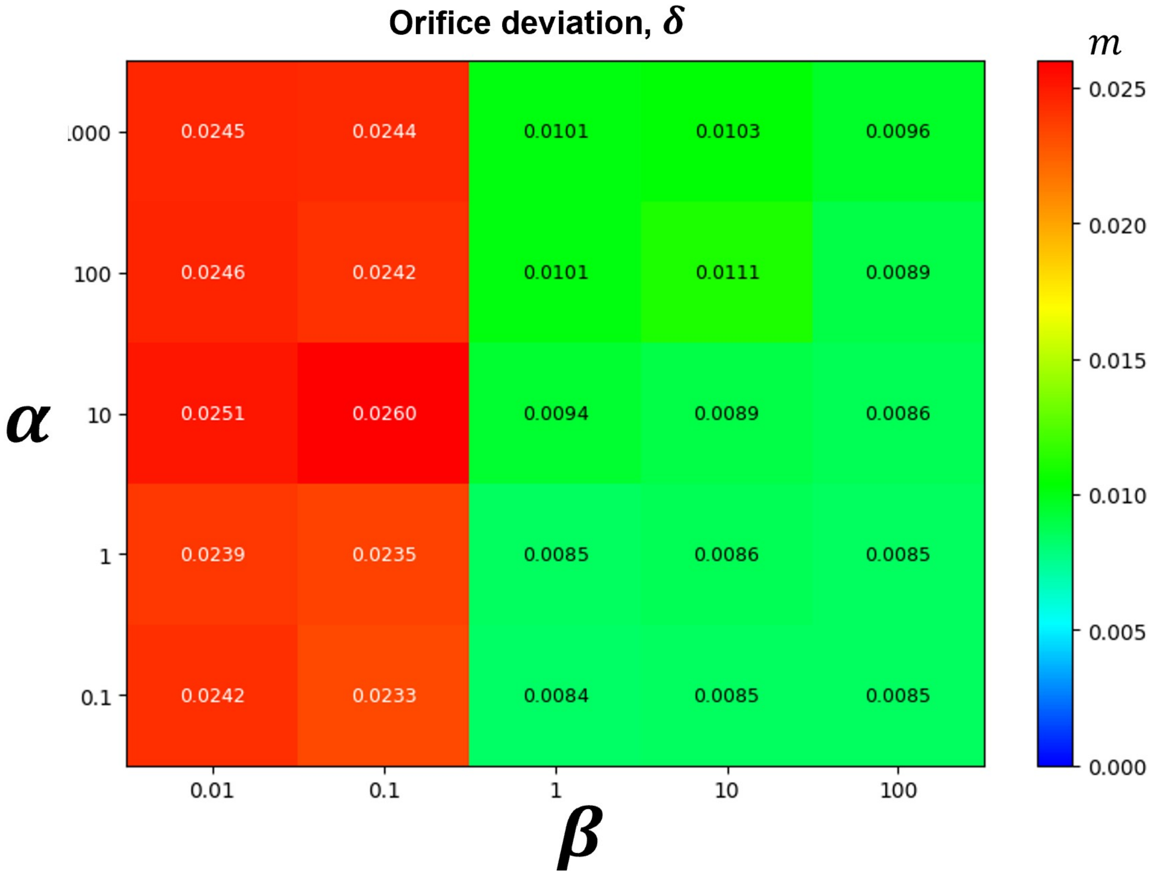


**Figure S8: Influence of Stiffness $\alpha$ and Landmark $\beta$ Parameters on Non-Rigid Registration.** The heatmap illustrates mean orifice deviation $\delta$ between IVUS and CT branch centroid locations as a function of $(\alpha, \beta)$. For each grid point, non-rigid registration was performed at the corresponding parameter pair across 3 patients (Patients 2, 4, and 6, validated against 4DCT), where the orifice deviation was averaged over all 4 abdominal branches per patient. The $5 \times 5$ grid yields a total of $N = 75$ non-rigid registrations.

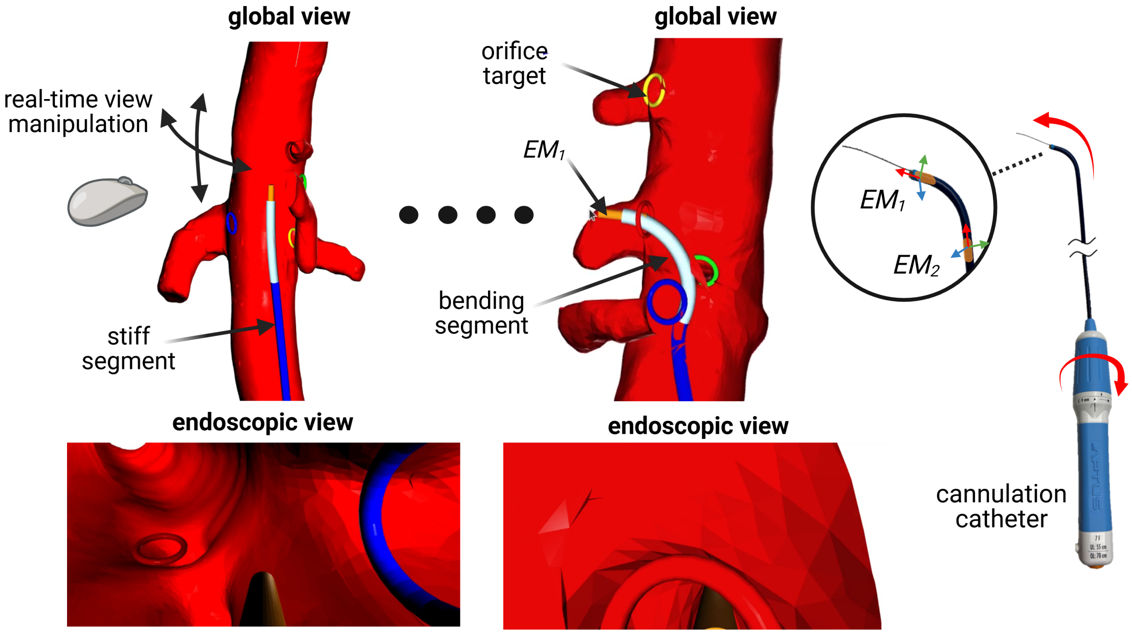


**Figure S9: Real-Time Augmented Reality Interface for Target Vessel Cannulation.** The interface provides dual complementary views: (**Top row**) Global 3D view showing the reconstructed aorta (red), catheter shaft (dark blue rigid segment, light blue steerable segment), and branch ostia (colored rings). The operator can interactively adjust the camera perspective in real-time. (**Bottom row**) Simulated endoscopic view with virtual camera at catheter tip, providing forward-looking visualization analogous to bronchoscopy. Branch targets appear as rings in the field of view, enabling intuitive "point-and-shoot" navigation. Two embedded EM trackers (base and tip) enable real-time catheter shape reconstruction and position tracking.

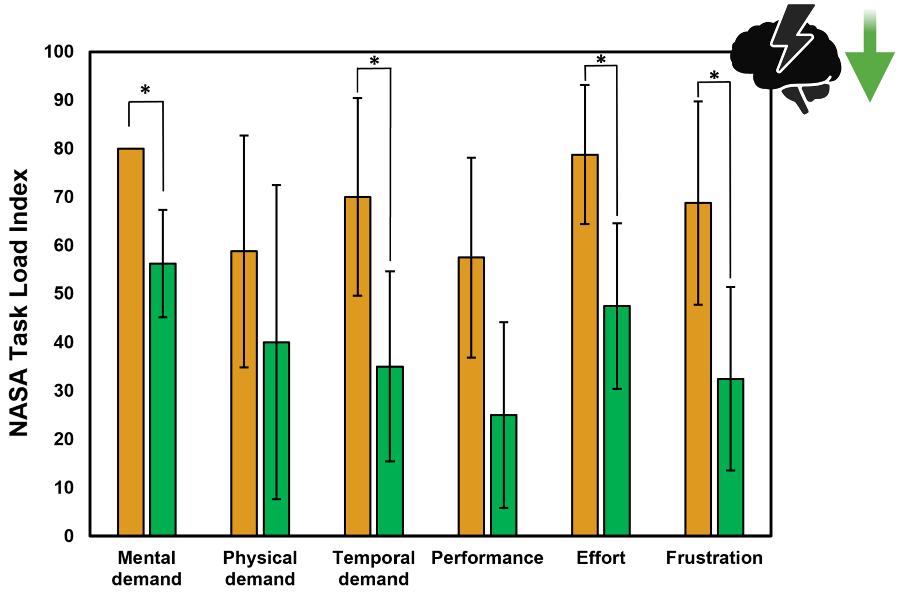


**Figure S10: NASA Task Load Index (TLX) results.** NASA Task Load Index scores across six cognitive dimensions. Lower scores indicate reduced workload in all cases.

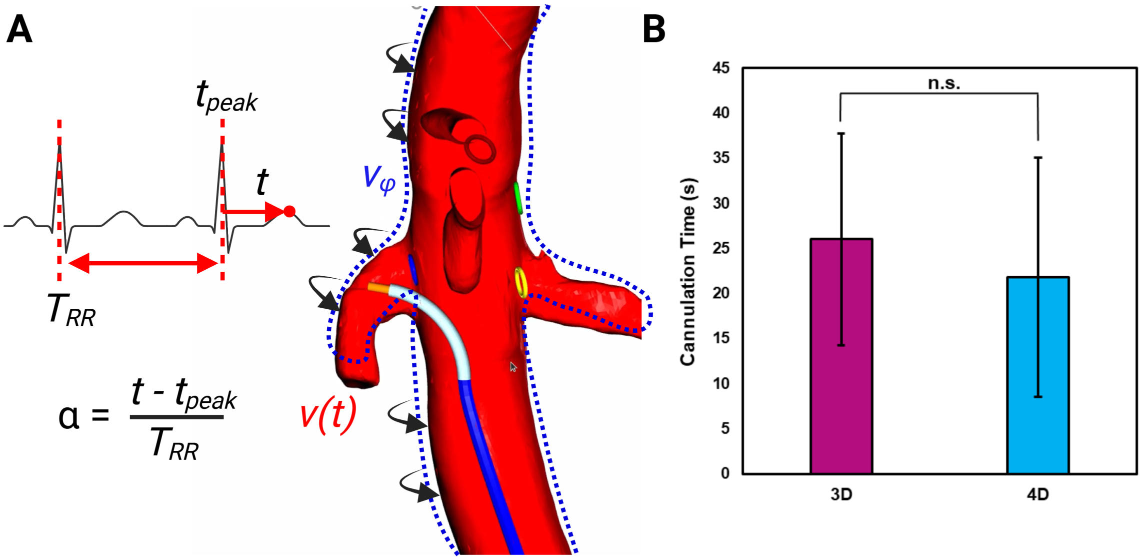


**Figure S11: Real-Time ECG-Synchronized Motion Compensation for Dynamic AR Guidance.** (**A**) The rendered CT mesh deforms in real time based on instantaneous cardiac phase ($\alpha$) computed from the ECG signal, tracking the phantom's cyclic motion throughout the cardiac cycle. Mesh vertex positions are interpolated between adjacent phase bins to provide smooth, continuous visualization. (**B**) Comparison of cannulation performance with 4D motion-compensated versus 3D static guidance across three patient-specific phantoms ($N = 48$ cannulations: 4 branches $\times$ 3 phantoms $\times$ 4 surgeons). Error bars represent standard deviation.

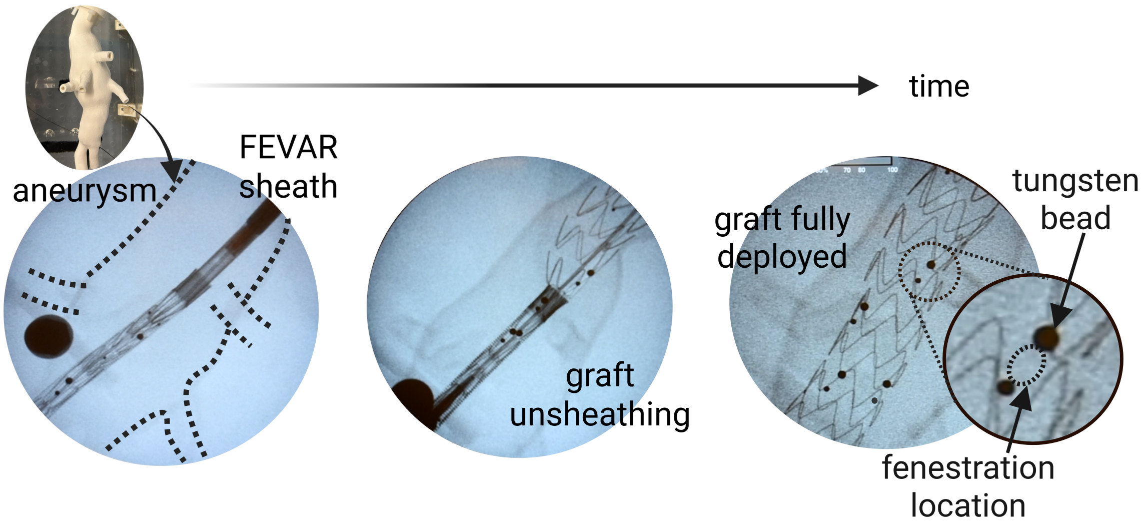


**Figure S12: Fenestrated Graft Deployment and IVUS-Based Fenestration Localization.** The fenestrated deployment system is introduced via femoral access and positioned within the aneurysm under fluoroscopic guidance. The graft is positioned rotationally and longitudinally to align the fenestrations to their corresponding branches. The fenestration boundaries (indicated by the dashed line) can be inferred from the spacing between adjacent tungsten beads.

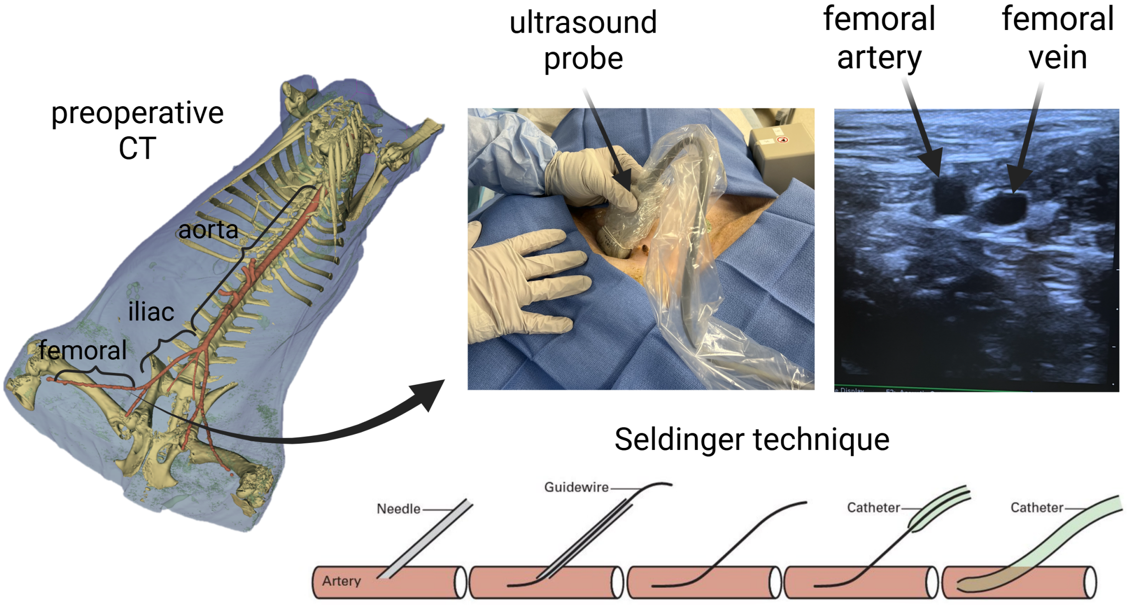


**Figure S13: Percutaneous Femoral Artery Access via Seldinger Technique.** Preoperative CT identifies the optimal femoral artery access site at the inguinal ligament level, and real-time ultrasound imaging can be used to idenitfy the femoral artery. The Seldinger technique is used to gain access, providing sealed vascular access for the hybrid IVUS-EM mapping catheter and steerable cannulation catheter.

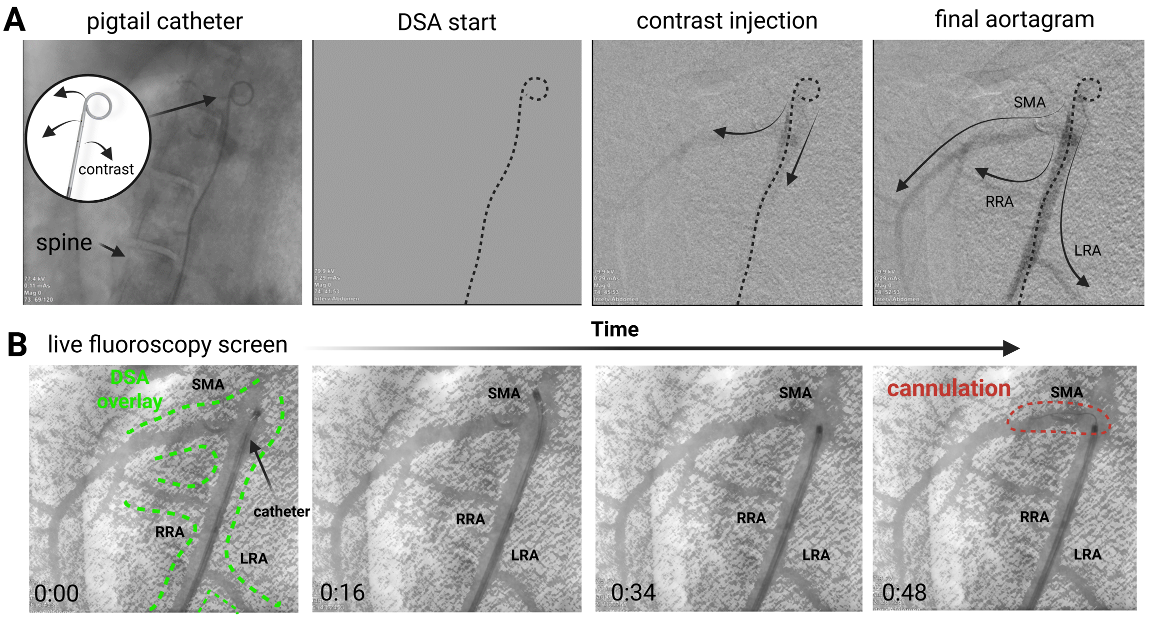


**Figure S14: Fluoroscopic Guidance *in vivo* with Digital Subtraction Angiography (DSA).** (**A**) A 5 Fr pigtail catheter positioned in the descending thoracic aorta delivers iodinated contrast into the abdominal aorta. The DSA acquisition visualizes major branch vessels. (**B**) The DSA roadmap is overlaid as a semi-transparent mask on live fluoroscopy, providing anatomical reference for target vessel cannulation.
